# Direct and indirect genetic pathways between parental neuroticism and offspring emotional problems across development: evidence from 7 cohorts across 5 European nations

**DOI:** 10.1101/2024.09.12.24313313

**Authors:** Hannah M. Sallis, Ilaria Costantini, Melisa T. Chuong, Katri Kantojärvi, Robyn E. Wootton, Hannah J. Jones, Lea Sirignano, Josef Frank, Fabian Streit, Stephanie H. Witt, Lea Zillich, Maria Gilles, Helga Ask, Alex S. F. Kwong, Mark J Adams, Kate Tilling, Deborah A. Lawlor, Nicholas J. Timpson, Tiina Paunio, Alexandra Havdahl, Andrew M. McIntosh, Alan Stein, Deborah James, Rebecca M. Pearson

## Abstract

Disentangling direct and indirect genetic pathways underlying the intergenerational transmission of emotional problems could guide preventative strategies and further the understanding of the role of parental mental health in children’s outcomes. This study aimed to estimate the extent to which genetic pathways that are *direct* (via child genotype) and *indirect* (e.g., via parental phenotype) explain the well-established association between parent and child emotional problems. We leveraged data from seven European cohort studies with a combined population of N*trios*=15,475. Polygenic scores were calculated for parental and offspring neuroticism, as it represents a dispositional trait underlying emotional problems. Emotional problems in offspring were measured using validated scales across various developmental stages from early childhood to adulthood. We used neuroticism polygenic scores within a structural equation modelling framework to distinguish between direct genetic pathways from parental genotype to offspring outcome (acting through offspring genotype), and indirect genetic pathways (acting through parental phenotype and associated environment). Standard errors for direct genetic, indirect genetic and total effects were bootstrapped and meta-analyses pooled effect estimates at three developmental stages (childhood: 3-4 years, adolescence: 11-13 years, adulthood: 18+ years). We found evidence suggesting an indirect genetic pathway between mothers and child emotional problems during early childhood (pooled estimate, mean difference in standardised child emotional problems score per 1SD increase in maternal PGS for neuroticism=0.04, 95% CI: 0.01, 0.07). This association attenuated over child development, while direct genetic pathways strengthened. High attrition rates, measurement error and low variance explained by polygenic scores may have altered precision of the estimates, influencing the interpretation of the results. However, we provide the first multi-cohort study to provide evidence for an *indirect* genetic pathway from maternal neuroticism to early child emotional problems. This suggests that there are likely processes *other* than direct genetic pathways involved in the intergenerational transmission of emotional problems, highlighting the importance of timely support to prevent and reduce emotional issues in mothers as a preventative strategy for emotional difficulties.

## Introduction

The prevalence of emotional problems, such as depression and anxiety(1), is increasing, particularly among young adults(2,3). Despite decades of research into ‘causal’ risk factors, policy changes, and intervention development, the burden of mental health disease has risen(4), with mental illness now being a leading cause of disability worldwide(5). Developing effective preventive interventions for emotional difficulties could have major societal and economic benefits by improving population health.

There is a well-established association between parent and child emotional problems. Family history of mental health problems for instance is considered one of the most important risk factors for the onset and maintenance of psychiatric disorders(6,7). In the UK, children with parents experiencing high levels of psychological distress are five times more likely to suffer from emotional disorders(8). This intergenerational risk is transmitted through both genetic and environmental mechanisms, as well as a likely interplay among the two. A key question is to identify potentially modifiable pathways to this intergenerational transmission of poor mental health, as different mechanisms of transmission will inform different preventative strategies.

Beyond an individuals’ genetic predisposition to a specific disorder (i.e., transmitted by the parents - direct genetic effect), non-transmitted parental genes can act as risk factors for the onset of emotional problems (also known as indirect genetic effect, “genetic nurture”, or familial effects(9)) by influencing aspects of the environment that are likely to contribute to child psychopathology and/or moderate the association between the child’s own genetic liability and mental health outcomes. Recent observational evidence and meta-analyses of genetically informed studies, including twin and adoption studies, have highlighted a clear role, albeit relatively small (12-20% of the variance), for the familiar environment in the development of psychopathology(10,11). A comprehensive review of genetically informative study designs, examining the association between parental and offspring emotional problems, reported strong evidence of both genetic and environmental transmission of parental depression to emotional problems across childhood, adolescence, and adulthood(12). Genetically informed studies can thus help to ascertain whether the associations between parental risk factors and offspring emotional outcomes are influenced by direct environmental effects or shared genetic factors. These designs offer improvements in accounting for genetic and shared environmental confounding factors compared to traditional observational studies(13,14).

Neuroticism is a personality trait indicating an individual’s tendency in experience negative emotions more frequently and intensely after exposure to stressors(15,16). It has also been conceptualised as a dispositional risk trait for emotional problems(16), including depression and anxiety, and its utility in research and clinical applications has been discussed elsewhere(17–19). Recent research shows that a genetic liability to neuroticism (proxied by a polygenic score – PGS) associates with emotional problems and depressive symptoms from infancy to young adulthood(20,21). Because neuroticism associates with both depression and anxiety, we decided to examine genetic liability to neuroticism as a common susceptibility to emotional problems. Using a genetic liability to a continuous personality trait as compared to a clinical disorder may be most useful when investigating symptom levels within general populations(22,23).

Attempting to isolate the role of parental phenotype is important because if parents’ emotional difficulties influence offspring via non-genetic paths, timely treatment of parental mental health symptoms *alone* could reduce the risk in the child. However, if there is only evidence for direct genetic pathways, this could suggest that focusing solely on parental mental health might not be as effective in prevention of child mental health risk as those including the child or with a specific focus on child’s mental health.

The aim of the current study is to estimate the extent to which the association between parent and child emotional problems is influenced by indirect genetic pathways and direct genetic pathways(9). Indirect genetic pathways, such as those mediated through parental phenotype, offer an opportunity to distinguish pathways operating through parental characteristics, distinct from pathways involving direct transmission of genetic information to the child (Figure 1).

**Figure 1.**
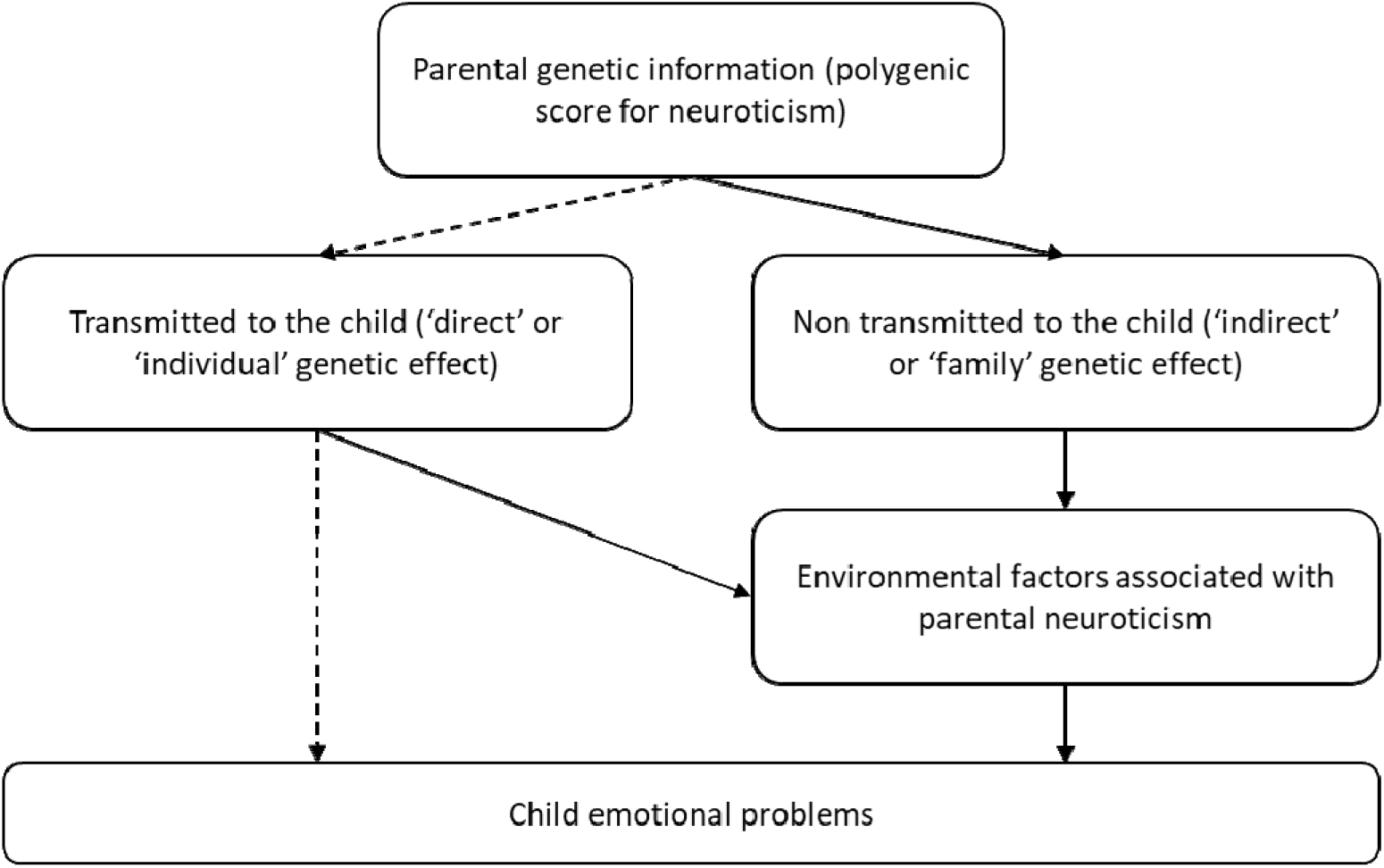
Potential pathways from genetic risk for neuroticism to offspring. This figure This figure illustrates the pathways of parental genetic transmission of risk to the offspring. In the case of child emotional problems, parental neuroticism variants influence parental liability to experience negative emotions, which in turn may shape the child rearing environment (i.e., nurture), ultimately affecting their offspring’s emotional difficulties. Importantly, both transmitted and non-transmitted parental genetic variants can contribute to the *indirect* genetic effect, for example by influencing parenting behaviors influenced by differing levels of neuroticism. In contrast, only transmitted parental alleles can be imlicated in *direct* genetic effects. Direct genetic pathways are represented by arrows with dotted lines, while indirect genetic pathways are illustrated by arrows with solid lines.

**Figure 2.**
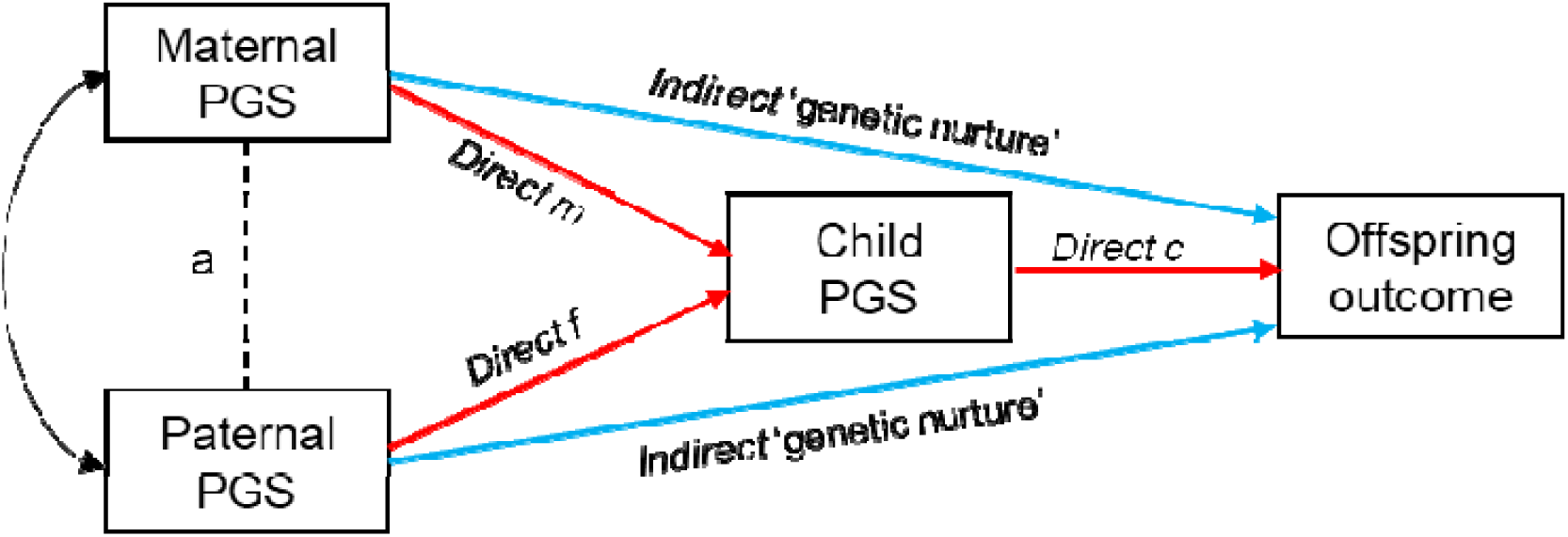
Example path model using trio genotype and offspring outcome measures. Relevant estimates can be obtained by regressing children’s outcomes onto their own polygenic scores as well as those of both parents. This multivariate trio model provides estimates of:

1. **Direct Genetic Effects**: These are the effects of parent-to-offspring allele transmission on the offspring’s outcome (red lines). This model adjusts for both maternal and paternal polygenic scores, thus isolating the direct genetic effects from the confounding genetic nurture effects. Specifically:

• **Direct m**: Maternal direct genetic effects (transmitted alleles to the offspring)
• **Direct f**: Paternal direct genetic effects (transmitted alleles to the offspring)
• **Direct c**: The pathway from the child’s own genetic liability to neuroticism to the phenotype of interest (e.g., offspring emotional problems)
2. **Indirect Genetic Effects**: These are the effects of parental alleles that have not been transmitted to the offspring (blue lines). Additionally, ‘a’ represents potential assortative mating between parents.

To address this aim, we used polygenic scores (PGS) for neuroticism, obtained from mother-father-offspring trios, and a pathway approach disentangling direct genetic and indirect genetic pathways to emotional disorders in offspring in childhood, adolescence, and adulthood (Figure 2). While previous studies using similar methods have identified indirect genetic pathways in educational outcomes(24), this study aims to extend our current understanding to mental health outcomes(25) by aggregating data from seven cohorts to enhance statistical power and to balance different sources of bias (i.e., different cohorts have different attrition patterns and reporting of emotional problems)(26).

Understanding the potential contribution of direct and indirect pathways may help guiding future strategies to prevent intergenerational cycles of emotional difficulties and improve child mental health in those who have a parent with emotional difficulties.

## Samples and methods

### Participants and genotyped data

This study used data from seven cohort studies across five countries, constituting the recently established Genetic and Environmental Mental health INtergenerational Investigation (GEMINI) consortium. We included all cohorts with both genetic and phenotypic data on the family trio. Detailed information for each cohort is provided in the Supplementary Information. In brief, this study combined information from the Avon Longitudinal Study of Parents and Children (ALSPAC)(27–29), Millenium Cohort Study (MCS)(30), Generation Scotland (GS)(31), the Norwegian Mother, Father and Child Cohort Study (MoBa)(32), FinnBrain (FB)(33), CHILD-SLEEP (CS)(34), and the Pre-, Peri-, and Postnatal Stress: Epigenetic Impact on Depression study (POSEIDON)(35,36). Please note that the ALSPAC website contains details of all the data that is available through a fully searchable data dictionary and variable search tool (http://www.bristol.ac.uk/alspac/researchers/our-data/).

## Measures

Offspring emotional problems and depressive symptoms were measured using different scales across development (validated for children’s ages) and reported by different assessors (i.e., parents and children – self-reported). Emotional problem scores were based on the following measures (STable 1) and standardised prior use in analyses to facilitate comparison and interpretability. Childhood emotional problems were measured using the Brief Infant-Toddler Social and Emotional Assessment (BITSEA)(37), the Child Behaviours Checklist (CBCL)(38), and Strengths and Difficulties Questionnaire (SDQ)(39). Adolescent emotional problems were measured using the Strengths and Difficulties Questionnaire (SDQ)(39) and the Short Moods and Feelings Questionnaire (SMFQ)(40). Finally, adult depressive symptoms were measured using The Clinical Interview Schedule-Revised (CIS- R)(41), General Health Questionnaire (GHQ)(42), and the Patient Health Questionnaire-2 (PHQ2)(43)

## Polygenic scores indexing parental and offspring neuroticism

Information on genotyping in each cohort is available in Supplementary Information. SNPs were filtered on minor allele frequency (>1%) and imputation info score (>0.8). In each cohort, PGS for neuroticism were derived based on publicly available summary statistics from a genome-wide association study by Luciano et al. (44). This study analysed over 329,000 individuals of self-described white British ancestry in the UK Biobank Study (46.3% male). Neuroticism was measured using a 12-item scale from the Eysenck Personality Questionnaire-Revised (EPQ-R). Neuroticism scores were adjusted for age, sex, genotyping batch, genotyping array, assessment centre and 40 principal components of ancestry prior to analysis. PGS were derived in each of the cohorts using PRSice2 with default clumping options (clumping correlated SNPs within a 250kb window at a r2 threshold of 0.1) and based on a p-value threshold of p<0.05. This threshold was chosen to maximise the variance explained by the PGS while minimising noise. All scores were standardised as z-scores prior to use in analyses. Betas (β) indicate a unit difference increase or reduction in the standardised outcome score for each 1SD increase in the exposure (i.e., PGS for neuroticism of child, mother, or father).

## Data analysis

### Descriptive analyses and missing data

We calculated medians and interquartile ranges (IQR) or frequencies and percentages for basic demographic characteristics of the participants across cohort studies and reported these data for mothers, fathers, children and trios with genetic data available (that were included in analyses because had the outcome), and those with missing genetic and outcome data (STable 2).

### Primary analyses

In each cohort we used neuroticism PGS from the mothers, fathers, and offspring within a SEM (Structural Equation Modelling) path analysis using Stata v16(45) (ALSPAC, MCS) or lavaan package in R (GS, MoBa, FinnBrain, ChildSleep, POSEIDON) (see supplementary file for exact coding) to separate out the direct genetic pathways of parental genotype (which acts via offspring genotype) on offspring emotional problems from the indirect genetic pathway that could act via maternal phenotype and environment (Figure 2). More specifically maternal and paternal PGS scores were regressed on child PGS scores and in turn child emotional outcomes and on child emotional outcomes directly. Because we included all pathways (both direct and indirect) from maternal and paternal PGS in a simultaneous model the path estimates allow us to deconstruct direct and indirect contributions. Among cohorts where population structure is more varied and thus more likely to introduce issues due to confounding by population stratification (e.g. GS, MCS), we accounted for 10 principal components of ancestry.

Standard errors were bootstrapped in each cohort and the effect estimates and 95% confidence intervals for the indirect and direct effects of maternal and paternal PRS were pooled in a random effects meta-analysis at three separate time points (childhood: age 2-4 years, adolescence: 11-13 years, adulthood: 18-99 years).

The measures were also combined according to rater (self-report or parent-report) across all time points in a random effects meta-analysis. This study follows STROBE reporting guidelines (STable 3).

## Results

### Samples descriptives

Sample demographic characteristics are summarised in Table 2. In ALSPAC, mothers with genetic data were more likely to be married or co-habiting, and were less likely to have ever smoked compared to mothers for whom genetic data was missing. In ALSPAC, MoBa, and GS, individuals with completed trio genetic data were more likely to have mothers with higher educational qualifications as compared to the full sample, while this is not the case for MCS(46,47). These descriptive results are summarised in STable 2 and reported in more details here(48).

Our pooled sample was 15,475 in the childhood analysis (aged 2-4 years), 4,507 in the adolescent analysis (11-13 years), and 4,244 in the adult analysis (18-99 years).

### Direct and indirect genetic effects on childhood emotional problems in six cohorts

In childhood, data on parent-reported emotional problems in childhood was available from 6 of the cohorts (i.e., ALSPAC, Child Sleep, FinnBrain, MCS, MoBa, Poseidon). Evidence was strongest for an association between the maternal indirect genetic association (0.04SD difference in emotional problems per 1 standard deviation (SD) in maternal neuroticism PGS (95% CI: 0.01, 0.07), p=0.018) and child emotional problems. The effect estimates for direct genetic associations and paternal indirect genetic associations were in the same direction, but evidence was weaker for these effects (Figure 3, STable 4). However, the CIs for each of the estimates overlapped. We found some evidence of both maternal and paternal total effects.

**Figure 3.**
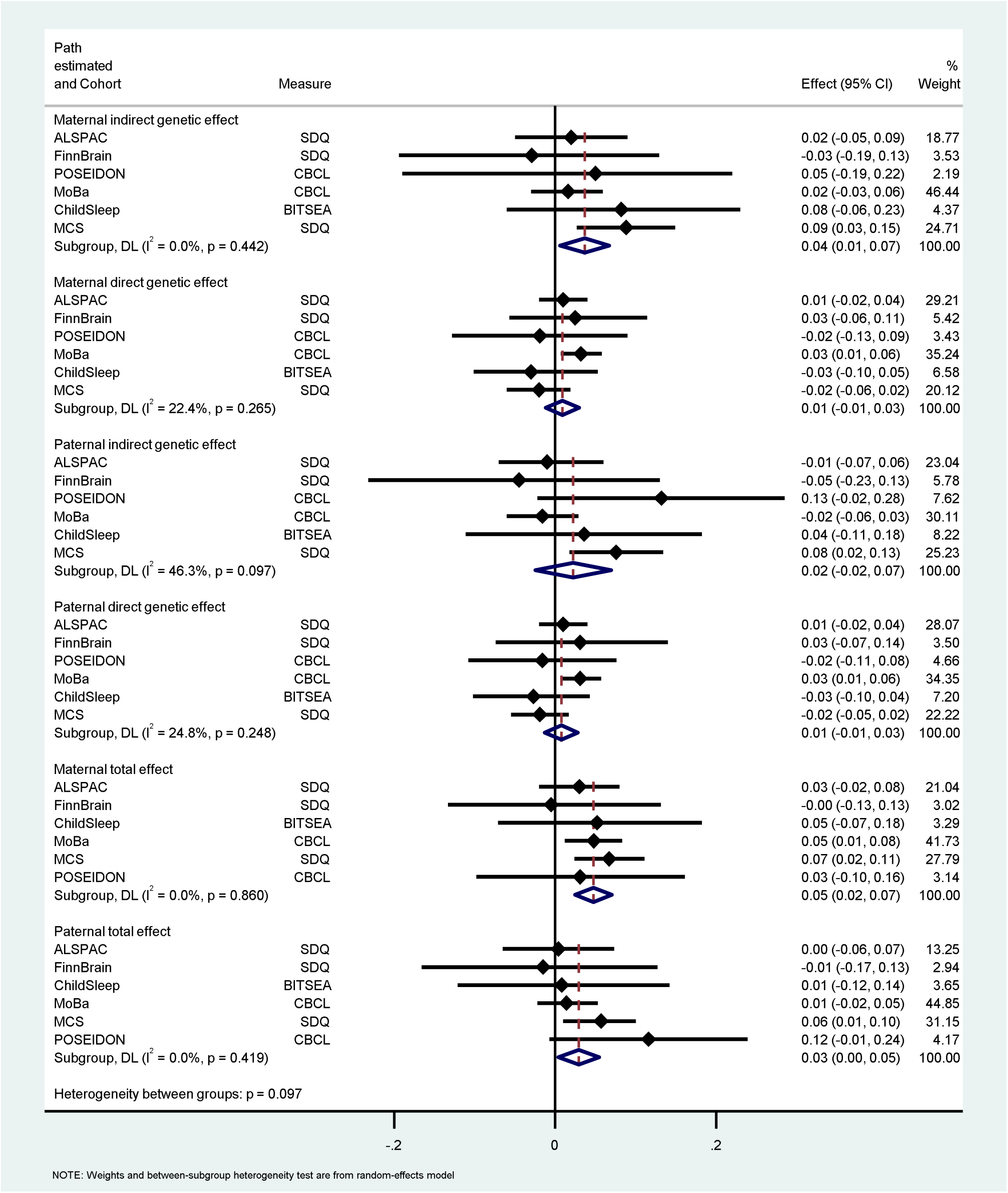
Association between parental genetics and childhood outcomes in ALSPAC, FinnBrain, ChildSleep, POSEIDON, MoBa, and MCS.

### Direct and indirect genetic effects on adolescent emotional problems and depressive symptoms in two cohorts

In adolescence, data on offspring emotional problems were available from ALSPAC and MCS cohorts (N=4,507). Here, we found some evidence of an association between both maternal and paternal direct genetic pathways and offspring emotional problems (Figure 4, STable 4). The direction of this association was consistent with that observed during childhood. For maternal direct genetic effects there was a 0.04SD difference in emotional problems per 1SD increase in maternal neuroticism PGS ((95% CI: 0.01, 0.06), p=0.010), while for paternal direct genetic effects there was a 0.03SD in difference in offspring emotional problems per 1SD increase in paternal neuroticism PGS (95% CI: 0.01, 0.06; p=0.006). While the direct genetic effect estimates are consistent between ALSPAC and MCS, the indirect genetic effect estimates appear to be in conflicting directions. In a similar way to early childhood, the CIs for all pathways overlap and the evidence for a total effect is less clear.

**Figure 4.**
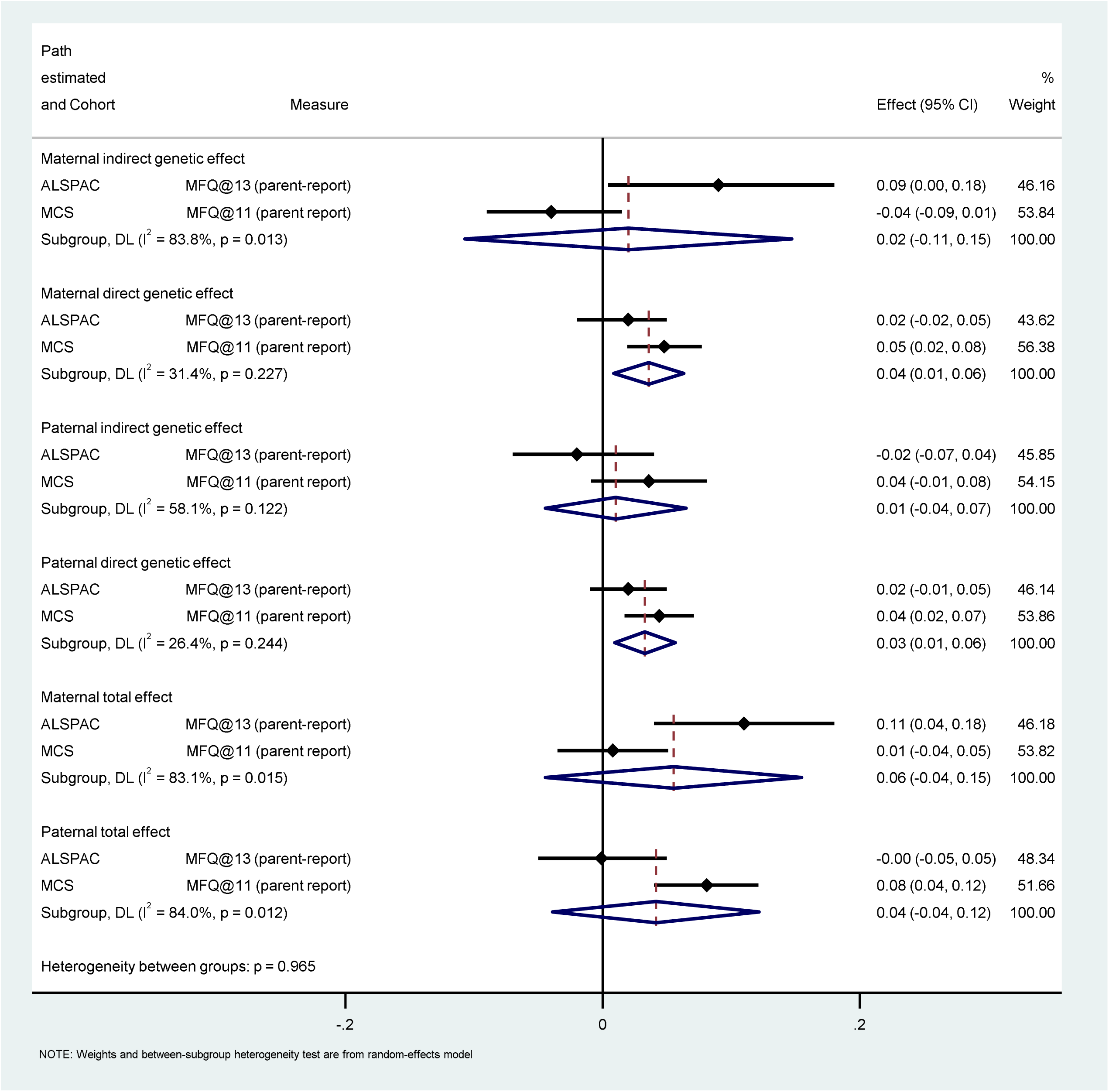
Effect of parental genetics on parent-reported adolescent outcomes in ALSPAC and MCS.

### Direct and indirect genetic effects on adulthood depressive symptoms in three cohorts

In adulthood, self-reported depressive symptoms data were available from ALSPAC, GS, and MCS cohorts (N=4,210). We found strong evidence for an association between both maternal and paternal direct genetic effects and offspring emotional problems (Figure 5, STable 4). For maternal direct genetic effects there was a 0.05SD difference in emotional problems per 1SD increase in maternal neuroticism PGS (95% CI: 0.02, 0.08; p<0.001), while for paternal direct genetic effects there was a 0.05SD in difference in offspring depressive symptoms per 1SD increase in paternal neuroticism PGS (95% CI: 0.02, 0.07; p<0.001). We found no evidence of an association between indirect genetic effects and offspring depressive symptoms. Confidence intervals, in particular for the maternal effects, were overlapping across pathways, and evidence of total effects was unclear, particularly for paternal effects on offspring depressive symptoms in adulthood.

**Figure 5.**
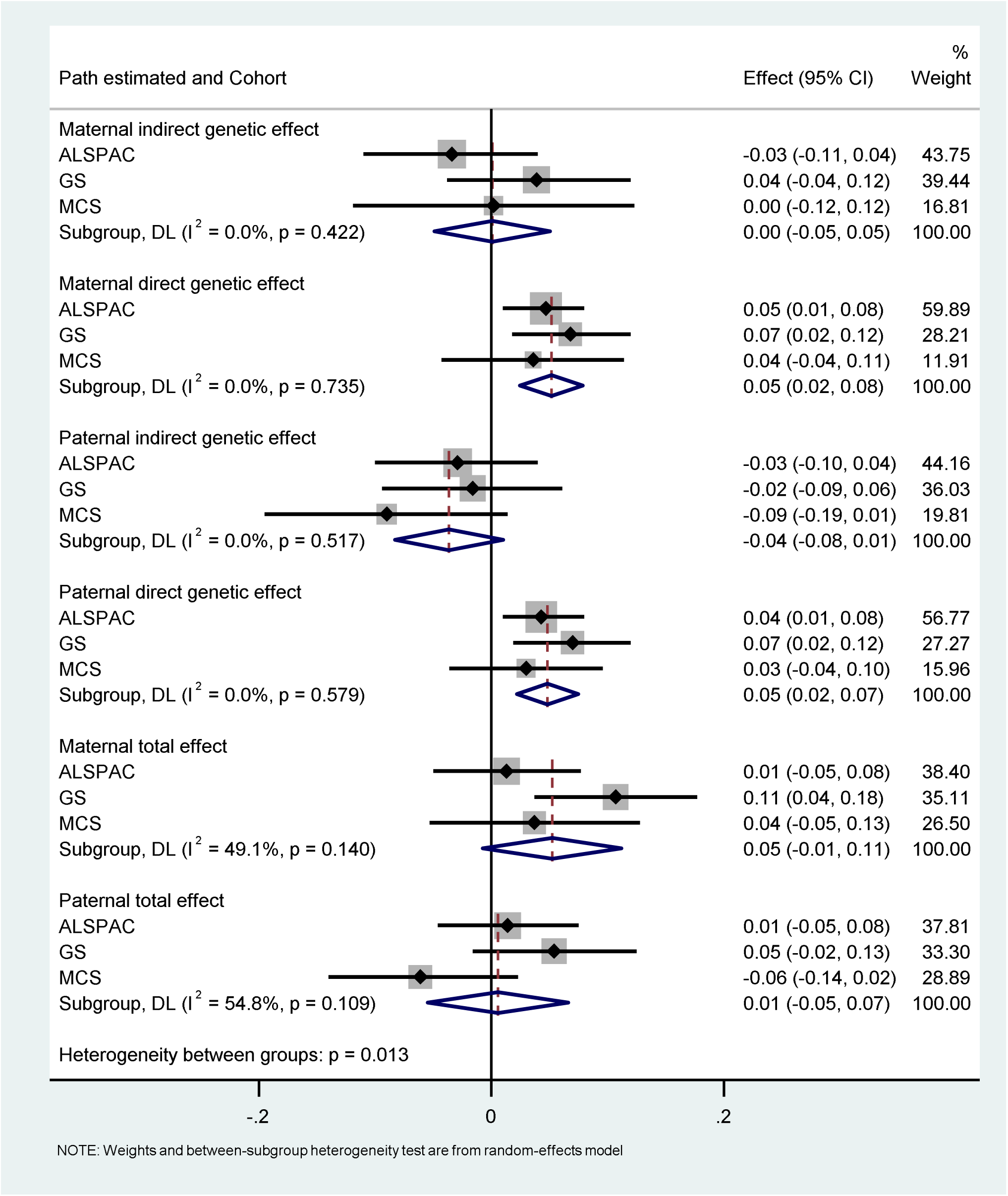
Effect of parental genetics on CIS-R depression score and the GHQ depression score in adulthood in ALSPAC, Generation Scotland, and MCS.

**Figure 6.**
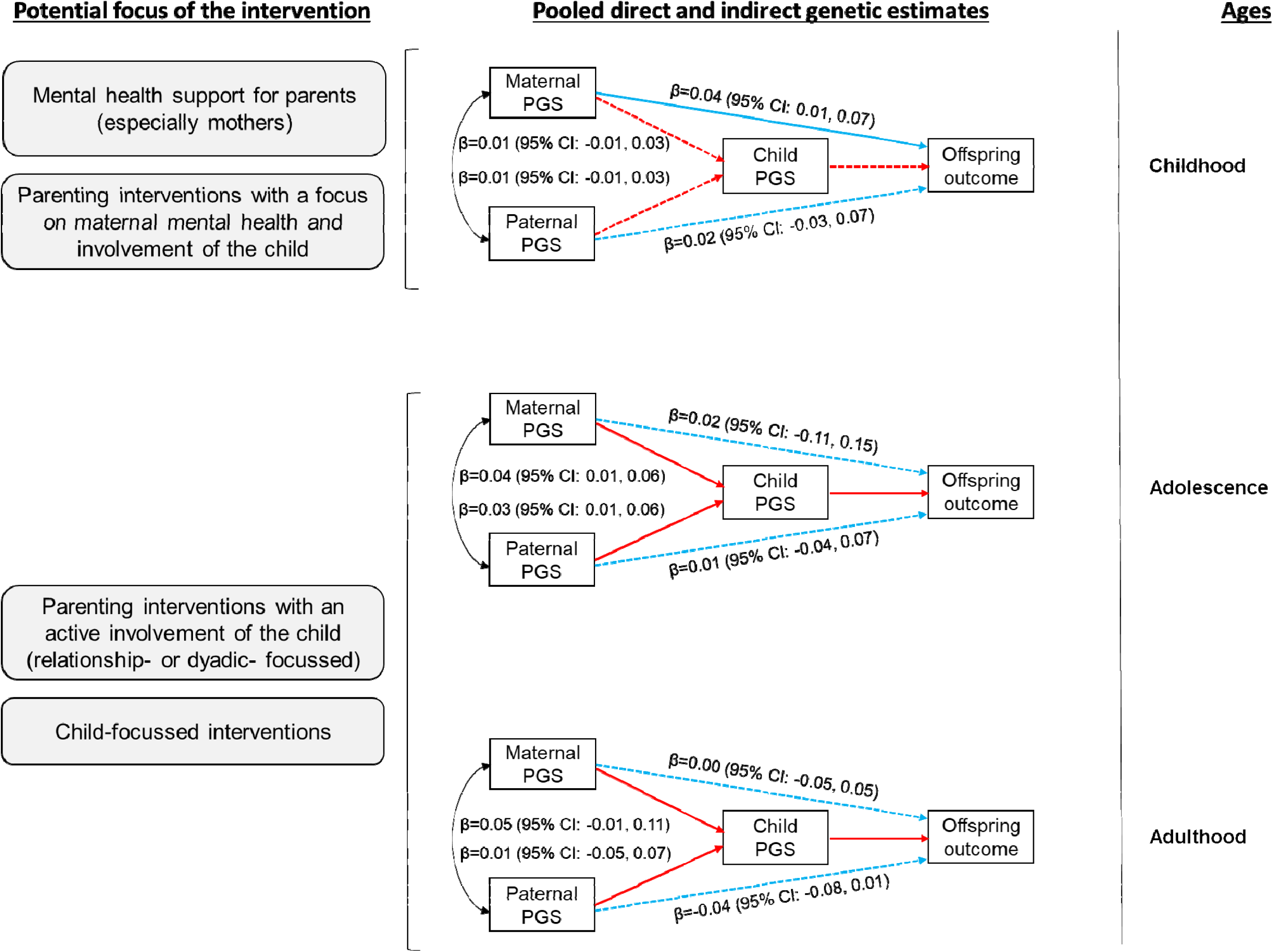
Pooled direct and indirect genetic effects across development and hypothesised intervention targets. Solid lines indicate the hypothesised pathways that, if targeted by interventions, may have more beneficial effects on the offspring emotional difficulties.

There were modest positive associations reflective of maternal and paternal potentially modifiable phenotypes and as well as both parent genetic inheritance. This was strongest for maternal phenotype (0.04SD difference in emotional problems per 1SD in maternal neuroticism PGS (95%CI: 0.01, 0.07)) and weakest in magnitude for maternal and paternal genetic inheritance (both 0.01 SD per 1SD PGS (-0.01, 0.03)). All pooled estimates were imprecise.

## Discussion

This study leveraged data from seven cohort studies across five countries to elucidate the association between parental genetic predisposition to neuroticism and child emotional problems and depressive symptoms through both direct and indirect genetic pathways. Using a genetically informed approach and integrated polygenic scores within a structural equation model, we disentangled the influence of direct and indirect genetic pathways on this association. We also investigated whether this association persisted from early childhood to adulthood by performing a meta-analysis of emotional problems in early childhood (age 2-4 years), adolescence (11-13 years), and adulthood (18 years and over).

Overall, our pooled results report suggestive evidence of an association between indirect genetic maternal pathways and child emotional problems during early childhood (n=15,475). The indirect maternal genetic effects diminished over time while direct genetic effects intensified across the life course (in more than 4,000 individuals). The strength of the evidence supporting an indirect genetic pathway drastically reduced in adolescence and adulthood, with the magnitude of the effect size halving from childhood to adolescence and from adolescence to adulthood for the maternal indirect effect, and with inconsistent direction of the effect estimate for the paternal indirect effect. Studies with genetics data, including twins and children-of-twin studies, have suggested that familiar influences and indirect genetic effects may play a greater role on offspring phenotypes in early ages as compared to adolescence and adulthood where the effect of the own genetic liability is more likely to contribute to the phenotype(49–52). These hypotheses are also supported by evidence genetic heritability for several traits, including mental health related traits, increases over the lifespan(11,53).

Alternatively, it is possible that we observed stronger evidence for an indirect pathway in early childhood because of the use of parent vs self-reported measures at these time points, which may have induced differential biases in the reporting child emotional symptoms associated to maternal mental health characteristics. Nevertheless, some of these biases have been explored in other studies, even in ALSPAC, reporting often negligible effects(20,54) (see in limitations).

Unlike the direct genetic effects which show a consistent direction of association, the effect estimates for the indirect genetic pathway also appear to be in conflicting directions across ALSPAC and MCS in adolescence. The variability of the indirect pathway could suggest that the environment captured by the indirect genetic component is likely to differ across studies (e.g., ALSPAC participants were recruited a decade earlier and from a select region of the UK, while MCS was sampled from across the entire UK), thus being explained by differences specific to each cohort (e.g., measures, drop-out rates, selection processes).

### The context within the literature

The estimates for direct genetic pathways become stronger in magnitude and strength of evidence during adolescence and adulthood. This may be potentially due to a diminishing role of the parental environment, as captured by the genetic component, in shaping the offspring’s environment during these developmental stages. An alternative explanation could be due to the use of polygenic scores derived from adult GWAS summary statistics in our analysis. These scores may therefore explain more of the variation in the phenotype as cohort members reach adulthood(55). However, previous work has shown that even at younger ages the neuroticism polygenic scores are still associated with the phenotypes(20).

### Implications for intergenerational psychiatry

Our findings suggest that direct genetic pathways more strongly explain the associations between parental neuroticism and their children’s emotional difficulties in adolescence and adulthood. Contrarily, indirect genetic pathways, especially via maternal ‘genetic nurturing’, more strongly explain the association with child emotional problems in early childhood. These findings have separate potential implications for preventative interventions (Figure 6). In the case of direct genetic pathways, delivering preventative interventions exclusively targeting parental mental health is unlikely to have beneficial effects on child emotional problems in adolescence and adulthood. This, however, contrasts with trial evidence(56) that targeting parental mental health or parenting skills in parents with mental health disorders have modest effects in reducing internalising disorders in children regardless of their age (effects were observed in both childhood and adolescence). However, the magnitude of the effects is relatively small and interventions directly supporting child resilience, emotional regulation or parenting interventions with active involvement of the child may be more effective in reducing emotional problems. Such findings align though with meta-analyses of trials that found that parenting interventions with an active inclusion of the child and focus on the relationship are more effective in reducing internalising problems as interventions that involved parents only(57,58).

In the case of genetic nurture, the effects of interventions targeting parental mental health (especially maternal) would be expected to have beneficial downstream effects on child emotional problems as experienced in early childhood.

### Limitations

This study may be subject to several limitations which should be considered when evaluating its findings. First, despite being the largest study of its kind, limitations persist due to sample size constraints. The need for genetic data from mothers, fathers, and offspring has restricted the available sample, as many studies only offer genetic data for parent-offspring pairs. However, combining data across multiple cohorts has maximised the sample size, especially in early childhood, where statistical power is greater as compared to the other timepoints. Nevertheless, missing genetic information data and severe attrition over time may have introduced bias into estimates, as those providing genetic information and remaining in the cohort were more likely to come from more socio-economically advantaged families, especially in ALSPAC, CHILD-SLEEP, MoBa, and GS (see STable 2). This pattern of missingness could have influenced our findings, most likely biasing them toward the null as it known that higher education moderates the effects of poor maternal mental health on offspring emotional and behavioural difficulties (47). Future studies should aim to employ methods such as parental genetic imputation (59), inverse probability weighting(61) or multiple imputation(62) to minimise these issues. In addition, we have used data from five European countries therefore limiting the generalisability to this study to non-European participants.

Second, despite the increase in sample size of GWAS, they are still limited in estimating all the phenotypic variance explained by common variants, often resulting in these scores explaining only a small portion of the phenotypic variance. Consequently, when the estimates are used as weights in the construction of the polygenic score, they introduce noise(9). This noise can lead to observed null associations between the resulting polygenic scores and outcomes of interest, potentially reflecting false negatives where a true association exists but remains undetected.

Third, depression or emotional problems were primarily collected by a single reporter within each study and at a single time point which is likely to increase measurement error. Different reporters of internalising problems lead, for example, to different heritability estimates(63). To address potential reporting biases, having multiple raters assessing the phenotype would have been beneficial. However, this is challenging in childhood due to limited validation of child or teacher reports. Moreover, subjective ratings of mental health difficulties present inherent limitations regardless of the reporter. According to the ‘distortion model’(64–66), the mental health characteristics of the reporters may systematically bias the ratings of mental health questionnaires, with implications for assessing causality in genetic studies (9). However, previous investigations into such biases have found limited support for dependent differential misclassification or measurement of outcomes (20,54).

Finally, we cannot rule out possible bias due to assortative mating or population stratification, even though we have likely accounted for the latter by including a large number of principal components of genetic ancestry.

### Conclusions and direction for future research

In conclusion, our findings suggest that indirect genetic processes, such as the environment provided by the parent, are most strongly associated with offspring mental health in early life. Conversely, pathway estimates for directly transmitted genetic information become larger during adolescence and adulthood, although the individual reporting on emotional problems may play a role in this. This insight holds potential for refining intervention strategies aimed at mitigating the impact on offspring mental health and importantly the potential role of including *both* parent and child from as early as possible. The magnitude of the association was larger for the maternal indirect pathways as compared to the paternal indirect genetic pathways which may indicate the degree of parental involvement. Indeed, variation in parental involvement in children’s upbringing, such as time spent at home and involvement after separation, may also moderate the effects of indirect genetic effects on offspring phenotype with potentially more pronounced effects for the more involved parents. However, actual parental involvement was not explored in our study, and we recommend this for future investigations.

## Data Availability

Data from each individual cohort can be obtained through individual data request and will vary depending on the agreement with each cohort study. For example, ALSPAC data access is through a system of managed open access. Information on how to request access to ALSPAC data can be found on the ALSPAC website (http://www.bristol.ac.uk/alspac/researchers/access/).
Access to data sets requires approval from The Regional Committee for Medical and Health Research Ethics in Norway and an agreement with MoBa. For the MCS, data access is free of charge and it is detailed here https://cls.ucl.ac.uk/data-access-training/data-access/.

## Acknowledgments

We are extremely grateful to all the families who took part in this study, the midwives for their help in recruiting them, and the whole ALSPAC team, which includes interviewers, computer and laboratory technicians, clerical workers, research scientists, volunteers, managers, receptionists, and nurses. The Norwegian Mother, Father and Child Cohort Study is supported by the Norwegian Ministry of Health and Care Services and the Ministry of Education and Research. We are grateful to all the participating families in Norway who take part in this on-going cohort study. We thank the Norwegian Institute of Public Health (NIPH) for generating high-quality genomic data. This research is part of the HARVEST collaboration, supported by the Research Council of Norway (NRC) (#229624). We also thank the NORMENT Centre for providing genotype data, funded by NRC (#223273), South East Norway Health Authority and KG Jebsen Stiftelsen. Further we thank the Center for Diabetes Research, the University of Bergen for providing genotype data and performing quality control and imputation of the data funded by the ERC AdG project SELECTionPREDISPOSED, Stiftelsen Kristian Gerhard Jebsen, Trond Mohn Foundation, NRC, the Novo Nordisk Foundation, the University of Bergen, and the Western Norway Health Authorities (Helse Vest). We thank all parents and children for taking part in the POSEIDON study. We thank our student employees and interns for their support with data acquisition and data entry. We are grateful to the GS:SHFS participants in their families. Analysis of GS:SHFS data made use of the resources provided by the Edinburgh Compute and Data Facility (ECDF) (http://www.ecdf.ed.ac.uk/).

## Fundings

The UK Medical Research Council and Wellcome (Grant ref: 217065/Z/19/Z) and the University of Bristol provide core support for ALSPAC. This work was supported by European Research Council (ERC) under the European Union’s Seventh Framework Programme (grant FP/2007-2013), European Research Council Grant Agreements (grants 758813; MHINT). HS, RW, DAL, NJT work in or are affiliated with a unit that receives funding from the University of Bristol and UK Medical Research Council (MC_UU_00032/05, MC_UU_00032/06 and MC_UU_00032/07). DAL is contribution to this study is supported by the European Research Council (Grant Agreement 101021566 ART-Health), GWAS data was generated by Sample Logistics and Genotyping Facilities at Wellcome Sanger Institute and LabCorp (Laboratory Corporation of America) using support from 23andMe. A comprehensive list of grants funding is available on the ALSPAC website (http://www.bristol.ac.uk/alspac/external/documents/grant-acknowledgments.pdf). A. S. F. Kwong is supported by a Wellcome Early Career Award (Grant ref: 227063/Z/23/Z).

Generation Scotland: Scottish Family Health Study was supported by the Chief Scientist Office (CZD/16/6) and the Scottish Funding Council (HR03006) and by the Wellcome Trust (104036/Z/14/Z).

The POSEIDON study was supported by a grant of the Dietmar Hopp Foundation.

The Norwegian Mother, Father and Child Cohort Study is supported by the Norwegian Ministry of Health and Care Services, and the Ministry of Education and Research. A.H. is supported by the Research Council of Norway (#274611, #336085 and #288083), the South- Eastern Norway Regional Health Authority (#2020022), and her contribution to this study is supported by the European Union’s Horizon Europe Research and Innovation Programme (FAMILY, grant agreement No 101057529). Views and opinions expressed are however those of the author(s) only and do not necessarily reflect those of the European Union, or the other funding agents. Neither the European Union nor the granting authorities can be held responsible for them.

## Author contributions

This publication is the work of the authors and RMP, IC, and HS will serve as guarantors for the contents of this paper.

## Notes

### Competing Interest Statement

The authors have declared no competing interest.

### Author Declarations

Each cohort received its own ethic approval to conduct the study. For example, ALSPAC study was approved by the ALSPAC Ethics and Law Committee and the Local Research Ethics Committees. European Research Council Grant Agreements (grant: MHINT) provided the ethic approval for combining the cohorts.

## References

1. Durbeej N, Sörman K, Norén Selinus E, Lundström S, Lichtenstein P, Hellner C, et al. Trends in childhood and adolescent internalizing symptoms: results from Swedish population based twin cohorts. BMC Psychol. 2019;7(1):1–10.

2. Twenge JM, Cooper AB, Joiner TE, Duffy ME, Binau SG. Age, period, and cohort trends in mood disorder indicators and suicide-related outcomes in a nationally representative dataset, 2005-2017. J Abnorm Psychol. 2019 Apr;128(3):185–99.

3. Gunnell D, Kidger J, Elvidge H. Adolescent mental health in crisis. Vol. 361, BMJ (Clinical research ed.). England; 2018. p. k2608.

4. Gore FM, Bloem PJN, Patton GC, Ferguson J, Joseph V, Coffey C, et al. Global burden of disease in young people aged 10-24 years: a systematic analysis. Lancet. 2011 Jun;377(9783):2093–102.

5. McManus S, Meltzer H, Brugha TS, Beddington PE, Jenkins R. Adult Psychiatric Morbidity in England - 2007, Results of a household survey. NHS Digital. 2007;

6. Goodman SH. Intergenerational Transmission of Depression. Annu Rev Clin Psychol [Internet]. 2020 May 7;16(1):213–38. Available from: 10.1146/annurev-clinpsy-071519-113915

7. Costantini I. Childhood internalising problems: a multi-method approach to explore intergenerational pathways. 2023.

8. Newlove-Delgado T, F M, T W, D M, J D, S M, et al. Mental Health of Children and Young People in England, 2022. Leeds; 2022.

9. Pingault J, Allegrini AG, Odigie T, Frach L, Baldwin JR, Rijsdijk F, et al. Research Review: How to interpret associations between polygenic scores, environmental risks, and phenotypes. Journal of Child Psychology and Psychiatry [Internet]. 2022 Oct 28;63(10):1125–39. Available from: https://acamh.onlinelibrary.wiley.com/doi/10.1111/jcpp.13607

10. Nikstat A, Riemann R. On the etiology of internalizing and externalizing problem behavior: A twin-family study. PLoS One. 2020;15(3):e0230626.

11. Polderman TJC, Benyamin B, De Leeuw CA, Sullivan PF, Van Bochoven A, Visscher PM, et al. Meta-analysis of the heritability of human traits based on fifty years of twin studies. Nat Genet. 2015;47(7):702.

12. Jami ES, Hammerschlag AR, Bartels M, Middeldorp CM. Parental characteristics and offspring mental health and related outcomes: a systematic review of genetically informative literature. Transl Psychiatry [Internet]. 2021;11(1):197. Available from: 10.1038/s41398-021-01300-2

13. Harold GT, Leve LD, Sellers R. How Can Genetically Informed Research Help Inform the Next Generation of Interparental and Parenting Interventions? Child Dev [Internet]. 2017 Mar;88(2):446–58. Available from: https://onlinelibrary.wiley.com/doi/10.1111/cdev.12742

14. Pingault JB, O’Reilly PF, Schoeler T, Ploubidis GB, Rijsdijk F, Dudbridge F. Using genetic data to strengthen causal inference in observational research. Nat Rev Genet [Internet]. 2018 Sep 5;19(9):566–80. Available from: https://www.nature.com/articles/s41576-018-0020-3

15. Eysenck HJ. Neuroticism, Anxiety, and Depression. Psychol Inq [Internet]. 1991 Jan;2(1):75–6. Available from: http://www.tandfonline.com/doi/abs/10.1207/s15327965pli0201_17

16. Barlow DH, Curreri AJ, Woodard LS. Neuroticism and disorders of emotion: A new synthesis. Curr Dir Psychol Sci. 2021;30(5):410–7.

17. Lahey BB, Moore TM, Kaczkurkin AN, Zald DH. Hierarchical models of psychopathology: empirical support, implications, and remaining issues. World Psychiatry [Internet]. 2021 Feb 12;20(1):57–63. Available from: https://onlinelibrary.wiley.com/doi/10.1002/wps.20824

18. Lahey BB. Public health significance of neuroticism. Am Psychol [Internet]. 2009;64(4):241–56. Available from: https://pubmed.ncbi.nlm.nih.gov/19449983

19. Cuijpers P, Smit F, Penninx BWJH, de Graaf R, ten Have M, Beekman ATF. Economic costs of neuroticism: a population-based study. Arch Gen Psychiatry. 2010;67(10):1086–93.

20. Costantini I, Sallis H, Tilling K, Major-Smith D, Pearson RM, Kounali DZ. Childhood trajectories of internalising and externalising problems associated with a polygenic risk score for neuroticism in a UK birth cohort study. JCPP Advances [Internet]. 2023 Feb 22;n/a(n/a):e12141. Available from: 10.1002/jcv2.12141

21. Kwong ASF, Morris TT, Pearson RM, Timpson NJ, Rice F, Stergiakouli E, et al. Polygenic risk for depression, anxiety and neuroticism are associated with the severity and rate of change in depressive symptoms across adolescence. Journal of Child Psychology and Psychiatry. 2021;62(12):1462–74.

22. Waszczuk MA. The utility of hierarchical models of psychopathology in genetics and biomarker research. World Psychiatry [Internet]. 2021 Feb 12;20(1):65–6. Available from: https://onlinelibrary.wiley.com/doi/10.1002/wps.20811

23. Plomin R, Haworth CMA, Davis OSP. Common disorders are quantitative traits. Nat Rev Genet [Internet]. 2009;10(12):872–8. Available from: 10.1038/nrg2670

24. Wang B, Baldwin JR, Schoeler T, Cheesman R, Barkhuizen W, Dudbridge F, et al. Robust genetic nurture effects on education: A systematic review and meta-analysis based on 38,654 families across 8 cohorts. The American Journal of Human Genetics. 2021 Sep 2;108(9):1780–91.

25. Pingault JB, Barkhuizen W, Wang B, Hannigan LJ, Eilertsen EM, Corfield E, et al. Genetic nurture versus genetic transmission of risk for ADHD traits in the Norwegian Mother, Father and Child Cohort Study. Mol Psychiatry [Internet]. 2023;28(4):1731–8. Available from: 10.1038/s41380-022-01863-6

26. Shakeshaft A, Martin J, Dennison CA, Riglin L, Lewis CM, O’Donovan MC, et al. Estimating the impact of transmitted and non-transmitted psychiatric and neurodevelopmental polygenic scores on youth emotional problems. Mol Psychiatry [Internet]. 2023; Available from: 10.1038/s41380-023-02319-1

27. Boyd A, Golding J, Macleod J, Lawlor DA, Fraser A, Henderson J, et al. Cohort Profile: the ’children of the 90s’--the index offspring of the Avon Longitudinal Study of Parents and Children. Int J Epidemiol. 2013;42(1):111–27.

28. Fraser A, Macdonald-Wallis C, Tilling K, Boyd A, Golding J, Davey Smith G, et al. Cohort Profile: the Avon Longitudinal Study of Parents and Children: ALSPAC mothers cohort. Int J Epidemiol. 2013;42(1):97–110.

29. Northstone K, Lewcock M, Groom A, Boyd A, Macleod J, Timpson N, et al. The Avon Longitudinal Study of Parents and Children (ALSPAC): an update on the enrolled sample of index children in 2019. Wellcome Open Res [Internet]. 2019 Mar 14;4:51. Available from: https://wellcomeopenresearch.org/articles/4-51/v1

30. Connelly R, Platt L. Cohort profile: UK millennium Cohort study (MCS). Int J Epidemiol. 2014;43(6):1719–25.

31. Smith BH, Campbell A, Linksted P, Fitzpatrick B, Jackson C, Kerr SM, et al. Cohort Profile: Generation Scotland: Scottish Family Health Study (GS: SFHS). The study, its participants and their potential for genetic research on health and illness. Int J Epidemiol. 2013;42(3):689–700.

32. Magnus P, Birke C, Vejrup K, Haugan A, Alsaker E, Daltveit AK, et al. Cohort profile update: the Norwegian mother and child cohort study (MoBa). Int J Epidemiol. 2016;45(2):382–8.

33. Karlsson L, Tolvanen M, Scheinin NM, Uusitupa HM, Korja R, Ekholm E, et al. Cohort profile: the FinnBrain birth cohort study (FinnBrain). Int J Epidemiol. 2018;47(1):15– 16j.

34. Paavonen EJ, Saarenpää-Heikkilä O, Pölkki P, Kylliäinen A, Porkka-Heiskanen T, Paunio T. Maternal and paternal sleep during pregnancy in the Child-sleep birth cohort. Sleep Med. 2017;29:47–56.

35. Send TS, Gilles M, Codd V, Wolf I, Bardtke S, Streit F, et al. Telomere length in newborns is related to maternal stress during pregnancy. Neuropsychopharmacology. 2017;42(12):2407–13.

36. Nieratschker V, Massart R, Gilles M, Luoni A, Suderman MJ, Krumm B, et al. MORC1 exhibits cross-species differential methylation in association with early life stress as well as genome-wide association with MDD. Transl Psychiatry. 2014;4(8):e429–e429.

37. Briggs-Gowan MJ, Carter AS, Irwin JR, Wachtel K, Cicchetti D V. Brief Infant-Toddler Social and Emotional Assessment (BITSEA) mannual, version 2.0. 2002;

38. Achenbach TM, Rescorla LA. The Achenbach system of empirically based assessment (ASEBA) for ages 1.5 to 18 years. In: The use of psychological testing for treatment planning and outcomes assessment. Routledge; 2014. p. 179–214.

39. Goodman R, Ford T, Simmons H, Gatward R, Meltzer H. Using the Strengths and Difficulties Questionnaire (SDQ) to screen for child psychiatric disorders in a community sample. The British journal of psychiatry. 2000;177(6):534–9.

40. Turner N, Joinson C, Peters TJ, Wiles N, Lewis G. Validity of the Short Mood and Feelings Questionnaire in late adolescence. Psychol Assess. 2014;26(3):752.

41. Lewis G, Pelosi AJ. The manual of CIS-R. London: Institute of Psychiatry. 1992;

42. Goldberg P. The detection of psychiatric illness by questionnaire. Maudsley monograph. 1972;

43. Kroenke K, Spitzer RL, Williams JBW. The Patient Health Questionnaire-2. Med Care [Internet]. 2003 Nov;41(11):1284–92. Available from: https://journals.lww.com/00005650-200311000-00008

44. Luciano M, Hagenaars SP, Davies G, Hill WD, Clarke TK, Shirali M, et al. Association analysis in over 329,000 individuals identifies 116 independent variants influencing neuroticism. Nat Genet [Internet]. 2018 Jan 18;50(1):6–11. Available from: http://www.nature.com/articles/s41588-017-0013-8

45. StataCorp. College Station, TX: StataCorp LLC. 2019. Stata Statistical Software: Release 16.

46. Joshi H, Fitzsimons E. The UK Millennium Cohort Study: the making of a multi- purpose resource for social science and policy in the UK. Longit Life Course Stud. 2016;7(4):409–30.

47. Fitzsimons E, Moulton V, Hughes DA, Neaves S, Ho K, Hemani G, et al. Collection of genetic data at scale for a nationally representative population: the UK Millennium Cohort Study. Longit Life Course Stud. 2022;13(1):169–87.

48. Pearson RM, Braithwaite E, Cadman T, Culpin I, Costantini I, Cordero M, et al. To know and be known: the proportion of genetic similarity for liability for neuroticism in mother-child and mother-father dyads is associated with reported relationship quality. 2023;

49. Eley TC, McAdams TA, Rijsdijk F V, Lichtenstein P, Narusyte J, Reiss D, et al. The Intergenerational Transmission of Anxiety: A Children-of-Twins Study. American Journal of Psychiatry [Internet]. 2015 Apr 23;172(7):630–7. Available from: 10.1176/appi.ajp.2015.14070818

50. Briley DA, Tucker-Drob EM. Explaining the Increasing Heritability of Cognitive Ability Across Development. Psychol Sci [Internet]. 2013 Sep 1;24(9):1704–13. Available from: http://journals.sagepub.com/doi/10.1177/0956797613478618

51. Scourfield J, Rice F, Thapar A, Harold GT, Martin N, McGuffin P. Depressive symptoms in children and adolescents: changing aetiological influences with development. Journal of Child Psychology and Psychiatry [Internet]. 2003 Oct 8;44(7):968–76. Available from: https://acamh.onlinelibrary.wiley.com/doi/10.1111/1469-7610.00181

52. Ask H, Eilertsen EM, Gjerde LC, Hannigan LJ, Gustavson K, Havdahl A, et al. Intergenerational transmission of parental neuroticism to emotional problems in 8- year-old children: Genetic and environmental influences. JCPP Advances [Internet]. 2021 Dec 1;1(4):e12054. Available from: 10.1002/jcv2.12054

53. Bergen SE, Gardner CO, Kendler KS. Age-related changes in heritability of behavioral phenotypes over adolescence and young adulthood: a meta-analysis. Twin Research and Human Genetics. 2007;10(3):423–33.

54. Olino TM, Michelini G, Mennies RJ, Kotov R, Klein DN. Does maternal psychopathology bias reports of offspring symptoms? A study using moderated non-linear factor analysis. Journal of Child Psychology and Psychiatry. 2021;

55. Kandler C. Nature and Nurture in Personality Development. Curr Dir Psychol Sci [Internet]. 2012 Oct;21(5):290–6. Available from: http://journals.sagepub.com/doi/10.1177/0963721412452557

56. Lannes A, Bui E, Arnaud C, Raynaud JP, Revet A. Preventive interventions in offspring of parents with mental illness: a systematic review and meta-analysis of randomized controlled trials. Psychol Med [Internet]. 2021/08/26. 2021;51(14):2321–36. Available from: https://www.cambridge.org/core/article/preventive-interventions-in-offspring-of-parents-with-mental-illness-a-systematic-review-and-metaanalysis-of-randomized-controlled-trials/24D36882C376B505170BD1A0DABB0101

57. Costantini I, López-López JA, Caldwell D, Campbell A, Hadjipanayi V, Cantrell SJ, et al. Early parenting interventions to prevent internalising problems in children and adolescents: a global systematic review and network meta-analysis. BMJ Mental Health [Internet]. 2023 Oct 1;26(1):e300811. Available from: http://mentalhealth.bmj.com//content/26/1/e300811.abstract

58. Wyatt Kaminski J, Valle LA, Filene JH, Boyle CL. A Meta-analytic Review of Components Associated with Parent Training Program Effectiveness. J Abnorm Child Psychol [Internet]. 2008;36(4):567–89. Available from: 10.1007/s10802-007-9201-9

59. Pearson RM, Evans J, Kounali D, Lewis G, Heron J, Ramchandani PG, et al. Maternal Depression During Pregnancy and the Postnatal Period: Risks and Possible Mechanisms for Offspring Depression at Age 18 Years. JAMA Psychiatry [Internet]. 2013 Dec 1;70(12):1312–9. Available from: 10.1001/jamapsychiatry.2013.2163

60. Young AI, Nehzati SM, Benonisdottir S, Okbay A, Jayashankar H, Lee C, et al. Mendelian imputation of parental genotypes improves estimates of direct genetic effects. Nat Genet [Internet]. 2022;54(6):897–905. Available from: 10.1038/s41588-022-01085-0

61. Austin PC, Stuart EA. Moving towards best practice when using inverse probability of treatment weighting (IPTW) using the propensity score to estimate causal treatment effects in observational studies. Stat Med. 2015;34(28):3661–79.

62. Sterne JAC, White IR, Carlin JB, Spratt M, Royston P, Kenward MG, et al. Multiple imputation for missing data in epidemiological and clinical research: potential and pitfalls. Bmj. 2009;338:b2393.

63. Jami ES, Hammerschlag AR, Ip HF, Allegrini AG, Benyamin B, Border R, et al. Genome-wide Association Meta-analysis of Childhood and Adolescent Internalizing Symptoms. J Am Acad Child Adolesc Psychiatry [Internet]. 2022;61(7):934–45. Available from: https://www.sciencedirect.com/science/article/pii/S0890856722001794

64. Vierhaus M, Lohaus A. Children and parents as informants of emotional and behavioural problems predicting female and male adolescent risk behaviour: A longitudinal cross-informant study. J Youth Adolesc. 2008;37(2):211.

65. Fergusson DM, Lynskey MT, Horwood LJ. The effect of maternal depression on maternal ratings of child behavior. J Abnorm Child Psychol [Internet]. 1993;21(3):245–69. Available from: 10.1007/BF00917534

66. Müller JM, Achtergarde S, Furniss T. The influence of maternal psychopathology on ratings of child psychiatric symptoms: an SEM analysis on cross-informant agreement. Eur Child Adolesc Psychiatry [Internet]. 2011;20(5):241–52. Available from: 10.1007/s00787-011-0168-2

