## Supplementary Material for "Direct and indirect genetic pathways between parental neuroticism and offspring emotional problems across development: evidence from 7 cohorts across 5 European nations"

### **Supplementary information**

#### **Cohort details**

##### **Avon Longitudinal Study of Parents and Children (ALSPAC)**

ALSPAC is a longitudinal pregnancy cohort which aimed to recruit all pregnant women in the former county of Avon with an expected due date between April 1991 and December 1992. Detailed information has continued to be collected on mothers, partners and children in the cohort, this process has been described in detail elsewhere ^1–3^. Ethics approval for the study was obtained from the ALSPAC Ethics and Law Committee and the Local Research Ethics Committees. Informed consent for the use of data collected via questionnaires and clinics was obtained from participants following the recommendations of the ALSPAC Ethics and Law Committee at the time. Please note that the study website contains details of the data that is available through a fully searchable data dictionary and variable search tool <https://variables.alspac.bris.ac.uk/>.

###### **ALSPAC genetic data generation and quality control**

ALSPAC children were genotyped using the Illumina HumanHap550 quad chip genotyping platforms. The resulting raw genome-wide data were subjected to standard quality control methods. Individuals were excluded on the basis of gender mismatches; minimal or excessive heterozygosity; disproportionate levels of individual missingness (>3%) and insufficient sample replication (IBD < 0.8). Population stratification was assessed by multidimensional scaling analysis and compared with Hapmap II (release 22) European descent (CEU), Han Chinese, Japanese and Yoruba reference populations; all individuals with non-European ancestry were removed. SNPs with a minor allele frequency of < 1%, a call rate of < 95% or evidence for violations of Hardy-Weinberg equilibrium (P < 5E-7) were removed. Cryptic relatedness was measured as proportion of identity by descent (IBD > 0.1). Related subjects that passed all other quality control thresholds were retained during subsequent phasing and imputation. 9,115 subjects and 500,527 SNPs passed these quality control filters.

ALSPAC mothers were genotyped using the Illumina human660W-quad array at Centre National de Génotypage (CNG) and genotypes were called with Illumina GenomeStudio. PLINK (v1.07) was used to carry out quality control measures on an initial set of 10,015 subjects and 557,124 directly genotyped SNPs. SNPs were removed if they displayed more than 5% missingness or a Hardy-Weinberg equilibrium P value of less than 1.0e-06. Additionally, SNPs with a minor allele frequency of less than 1% were removed. Samples were excluded if they displayed more than 5% missingness, had indeterminate X chromosome heterozygosity or extreme autosomal heterozygosity. Samples showing evidence of population stratification were identified by multidimensional scaling of genome-wide identity by state pairwise distances using the four HapMap populations as a reference, and then excluded. Cryptic relatedness was assessed using a IBD estimate of more than 0.125 which is expected to correspond to roughly 12.5% alleles shared IBD or a relatedness at the first cousin level. Related subjects that passed all other quality control thresholds were retained during subsequent phasing and imputation. 9,048 subjects and 526,688 SNPs passed these quality control filters.

After combining genotype data in the mothers and the children, SNPs with genotype missingness above 1% were removed due to poor quality (11,396 SNPs removed) and a further 321 subjects were removed due to potential ID mismatches. This resulted in a dataset of 17,842 subjects. Imputation of the target data was performed using Impute V2.2.2 against the 1000 genomes reference panel (Phase 1, Version 3) (all polymorphic SNPs excluding singletons), using all 2186 reference haplotypes (including non-Europeans).

This gave 8,237 eligible children and 8,196 eligible mothers with available genotype data after exclusion of related subjects using cryptic relatedness measures described previously.

3,453 ALSPAC mother and fathers and 535,478 SNPs were genotyped using the Illumina HumanCoreExome chip genotyping platforms by the ALSPAC lab and called using GenomeStudio. The resulting raw genome-wide data were subjected to standard quality control methods using PLINK (v1.07). Individuals were excluded on the basis of gender mismatches (n = 80); minimal or excessive heterozygosity (n = 64); disproportionate levels of individual missingness (>5%, n = 60) and possible contamination (n = 3). Population stratification was assessed by multidimensional scaling analysis and compared with 1000 Genomes phase 3 data and principal component analysis (n = 266); all individuals with non-European ancestry were removed. Cryptic relatedness was measured as SNP relatedness in GCTA (relatedness > 0.1, n = 69 removed). SNPs with a call rate of < 95% or evidence for violations of Hardy-Weinberg equilibrium (P < 1E-7) and those which failed GenomeStudio

quality control measures were removed (n = 21,298). 6,594 duplicate SNPs were also removed.

Data was phased for 3074 samples that passed QC but contained related subjects in SHAPEIT v2.r837. The following were then removed: 155,336 monomorphic SNPs, 1033 markers not in 1000 genomes, 11,842 A/T or G/C SNPs and 10 duplicate sites to give 337,732 SNPs on chromosomes 1-23. Of the 329,363 markers on chromosomes 1-22, 298,742 overlapped the reference genome. These were imputed to the 1000 genomes phase 1 version 3 using the Michigan Imputation Server. 1722 eligible fathers remained after QC, exclusion of duplicate subjects and individuals who had withdrawal of consent.

##### **CHILD SLEEP and FinnBrain**

Information on both CHILD SLEEP and FinnBrain genotyping have been provided in more details here^4^.

##### **Generation Scotland (GS)**

GS:SFHS comprises 24,000 adults (18+) from 7,000 families recruited from the general population of Scotland 2006-2011. 90% of the cohort attended a clinic in Dundee, Glasgow or Aberdeen where samples were taken and various tests carried out. The remainder returned a postal questionnaire and saliva sample for DNA. The study’s main aim is to investigate the genetics of common diseases, with a particular focus on mental health.

Further details here^5^.

GWAS data was obtained using the Illumina OmniExpress array, and imputed using the Haplotype Research Consortium (HRC) dataset. Further details of methods here^6^.

There were 7159 participants in the sample with genotype data who had at least one parent in the sample who also had genotype data. Because participants were embedded in a multi-generational, multi-sibling pedigree structure, one member from each sibling group with full trio data available was selected for analysis (N = 2,231). Depression scores were measured using the depression subscale of the General Health Questionnaire which was administered during a clinical visit following recruitment.

Ethical approval for the original data collection was obtained from the Tayside Committee on Medical Research Ethics A (ref 05/S1401/89). Generation Scotland is currently approved as a Research Tissue Bank by the East of Scotland Research Ethics Service (ref 20/ES/0021).

##### **Millennium Cohort Study (MCS)**

For details of data collection and genotyping, see the published ^7,8^.

##### **Norwegian Mother, Father and Child Cohort Study (MoBa)**

MoBa is a prospective population-based pregnancy cohort study conducted by the Norwegian Institute of Public Health^9^. Participants were recruited from all over Norway from 1999-2008. In 40.6 % of the pregnancies, women consented to participate. The cohort now includes 114,500 children, 95,200 mothers and 75,200 fathers. Blood samples were obtained from both parents during pregnancy and from mothers and children (umbilical cord) at birth. The current study is based on version 12 of the quality-assured data files released for research in January 2019. The establishment and initial data collection in MoBa were previously based on a license from the Norwegian Data protection agency and approval from The Regional Committees for Medical and Health Research Ethics (REC); MoBa is now regulated by regulations related to the Norwegian Health Registry Act. The current analyses were approved by REC (reference number 2016/1702).

###### **MoBa genetic data generation and quality control**

The genotyping and quality control (QC) procedure is detailed here^10^.

PLINK version 1.90 beta 3.36 (http://pngu.mgh.harvard.edu/purcell/plink/) was used to conduct the quality control, which has previously been described by Helgeland et al. 2019^11^. Known problematic SNPs previously reported by the Cohorts for Heart and Aging Research in Genomic Epidemiology (CHARGE) consortium and Psychiatric Genomics Consortium (PGC) were excluded from each batch. Duplicate samples were removed, and each genotyping batch was split into parents and offspring. Quality control was then conducted by genotyping array in parents and offspring separately.

Individuals were excluded if they had a genotyping call rate below 95% or autosomal heterozygosity greater than four standard deviations from the sample mean. SNPs were excluded if they were ambiguous (A / T and C / G), had a genotyping call rate below 98%, minor allele frequency of less than 1%, or Hardy-Weinberg equilibrium P-value less than 1 × 10-6. Population stratification was assessed, using the HapMap phase 3 release 3 as a reference, by principal component analysis using EIGENSTRAT version 6.1.4. Visual inspection identified a homogenous population of European ethnicity and individuals of non-European ethnicity were removed. Individuals with a genotyping call rate below 98% or autosomal heterozygosity greater than four standard deviations from the sample mean were then removed. A sex check was done by assessing the sex declared in the pedigree with the genetic sex, which was imputed based on the heterozygosity of chromosome X. When sex discrepancies were identified, the individual was flagged. Relatedness was assessed by flagging one individual from each pairwise comparison of identity-by-descent with a pi-hat greater than 0.1.

The parents and offspring datasets were then merged into one dataset per genotyping batch; keeping only the SNPs that passed quality control in both datasets. All individuals passing the genotyping call rate and autosomal heterozygosity measures were included in the merged datasets. Therefore, the merged datasets included individuals previously excluded or flagged as a duplicate, ethnic outlier, having a sex discrepancy, or high level of relatedness. Concordance checks were then conducted on validated duplicates. Duplicate, tri-allelic and discordant (any discordance between the validated duplicates) SNPs were excluded. Individuals and SNPs with a genotyping call rate below 98% in the merged datasets were excluded. The duplicate sample that was removed before the start of the quality control was then excluded. Mendelian errors identified by the assessment of duos and trios were then recoded to missing. Insertions and deletions were also excluded.

After QC the Human Core Exome 12 batch comprised 20,231 individuals and 384,855 SNPs, the Human Core Exome 24 batch comprised 12,757 individuals and 396,189 SNPs, and the Global Screening Array batch comprised 17,742 individuals and 568,275 SNPs. Phasing was conducted using Shapeit 2 release 837 and the duoHMM approach was used to account for the pedigree structure. Imputation was conducted using the Haplotype reference consortium (HRC) release 1-1 as the genetic reference panel. The Sanger Imputation Server was used to perform the imputation with the Positional Burrows-Wheeler Transform (PBWT). The phasing and imputation were conducted separately for each genotyping batch.

##### **Pre-, Peri-, and Postnatal Stress: Epigenetic Impact on Depression study (POSEIDON)**

*Subjects and samples*

POSEIDON (Pre-, Peri-, and Postnatal Stress: (Epi-) genetic impact on Depression; POSEIDON) is a longitudinal study including four time points: during the third trimester of pregnancy (T1), a few days after childbirth (T2), six months postpartum (T3) and 45 months postpartum (T4). For genetic analysis, saliva samples of the parents were assessed at T1 and for the children EDTA cord blood at birth (T2). The cohort included n=101 additional children and their parents at T4 to compensate for drop-out at T4. Here, salvia samples for the parents and the children were collected for genetic analysis. For participation in this meta-analysis, only complete trios (mother, father, child) with genetic and phenotypic data were included, resulting in a total n = 223 trios. The study took place following the Declaration of Helsinki and was approved by the Ethics Committee of the Medical Faculty Mannheim of the University of Heidelberg. All families provided written informed consent, which was (re)established at T4.

*Phenotype measurement*

The *internalising problems* subscale was measured at T4 based on ratings of the primary caregiver with the *German version of the Child Behavior Checklist for age from 1½ to 5 years* (CBCL)^12^. The *internalising problems* score was computed following the manual as the sum score of the 36 items from the subscale *internalising problems* and afterward z-standardized. Missings were replaced by the mean value of the available items of the scale for each individual (maximum 3 missing values per individual).

*Genotyping, Quality Control, and Imputation*

Genome-wide genotyping was performed using the Illumina Infinium Psych Array and the Illumina Global Screening Array (Illumina, Inc., San Diego, CA). Prior to imputation, the datasets were subjected a quality control (QC) procedure implemented in RICOPILI (Rapid Imputation and COmputational PIpeLIne for Genome-Wide Association Studies, from Broad Institute), including the following filtering steps: SNP missing rate < 0.05 (before filtering individuals), individual missingness < 0.02, exclusion of individuals with sex mismatches, autosomal heterozygosity deviation |F_het_| < 0.2, SNP missing rate < 0.02 (after filtering individuals), Minor Allele Frequency (MAF) > 0.01, Hardy–Weinberg Equilibrium (HWE) p ≥ 1e−06, and exclusion of SNPs without valid association p-value. Imputation was conducted with the RICOPILI imputation pipeline using the 1000 Genomes reference sample. After imputation, the two datasets (parents and children) were filtered with an additional SNP-missing rate < 0.02, INFO score ≥ 0.8, and merged resulting in an overlap SNP set of n=3,724,166. Relatedness check and principal component analysis (PCA) were conducted based on a quality filtered and pruned subset of autosomal SNPs (MAF > 0.2, pHWE > 0.02, SNP missingness < 0.0; pruning: pairwise R^2^ < 0.05 within a window of 250 SNPs). A relatedness cutoff of Pi Hat > 0.1 was used to exclude related individuals. To detect and remove genetic outliers an exceeding of six standard deviations on the first 20 PCA components was used. After QC, the dataset comprised n=277 complete trios from whom for n=223 the phenotype was available.

*Data availability*

Genetic and phenotypic data are not publicly available due to privacy regulations but are available from the authors on reasonable request.

#### **Measures**

All outcome variables were standardised as part of the model fitting procedure.

##### **Strenths and Difficulties Questioinnaire (SDQ)**

The Strengths and Difficulties Questionnaire (SDQ)^13^ is a brief behavioural screening questionnaire validated for children who are approximately 3–16 years old. Internalising scores are obtained by summing the emotional and the peer problems subscales and provides a score ranging from 0 to 20, where higher scores indicate a larger number of emotional difficulties exhibited by the child. ALSPAC and MCS measured the emotional difficulties of the child using the emotional problem subscale, whereas the CHILD SLEEP and FinnBrain used the internalising composite scale.

##### **Child Behaviours Checklist (CBCL)**

The Child Behavior Checklist (CBCL) was used to measure emotional problems (internalising items). The preschool checklist (CBCL/1½-5)^13^ version with 100 questions was used. The CBCL contains seven subscales in addition to a category of “other problems”. These are: Emotionally reactive, anxious/depressed, somatic complaints, withdrawn, sleep problems, attention problems and aggressive behaviour. The first four categories comprise a broader grouping of internalising symptoms. Higher scores indicate more emotional problems.

##### **Brief Infant-Toddler Social and Emotional Assessment (BITSEA)**

The BITSEA^14^ is a 31-item scale used to measure social–emotional/behavioral problems in 1- to 3-year-olds in a developmentally appropriate way. In this study we used the internalising scale which has 11 depression, anxiety, and negative emotionality items (e.g., “s*eems nervous, tense or fearful*").

##### **Short Mood and Feelings Questionnaire (SMFQ)**

The Short Mood and Feelings Questionnaire (SMFQ)^15^ is a 13-item questionnaire that measures the presence of depressive symptoms in the last two weeks. For each question, the answer can be “not true” (scored 0), “sometimes” (scored 1) and “true” (scored 2). As each question is scored between 0–2, the resulting summary score of all the items can range between 0–26, with higher scores being more indicative of greater depression.

##### **Clinical Interview Schedule – Revised (CIS-R): Depression scale**

Depression was measured using a self-administered, computerised version of the CIS-R completed during a study clinic attended when the participants were 17–18 years old.

**General Health Questionnaire: Depression Scale**

Mild psychological distress was assessed in GS:SHFS study using the 28-item General Health Questionnaire (GHQ-28)^16^ Depression was calculated as sum scores of the 7 items in subscale D, with the responses Not at all and “Not more than usual” scored as 0 and the responses “Rather more than usual” and “Much more than usual” scored as 1.

##### **Patient Health Questionnaire – 2 (PHQ2)**

The Patient Health Questionnaire (PHQ-2)^17^ includes the first two items of the PHQ-9. PHQ-2 was used to measure the frequency of two depressive symptoms (i.e., depressed mood: feeling down, depressed or hopeless; and anhedonia: little interest or pleasure in doing things) over the past 2 weeks. Each item was to be rated from 0 (“not at all”) to 3 (“nearly every day”).

#### **Supplementary Tables and Tables and Figures**

STable 1. Emotional Problems and Depressive Symptoms Measures Across Cohorts.

| **Measure** | **Subscale** | **Reporter** | **Age Means** | **Cohort** |
| --- | --- | --- | --- | --- |
| **Early Childhood** | | | |  |
| Brief Infant-Toddler Social and Emotional Assessment (BITSEA)^14^ | Internalising subscale | Parent | 2 years | ChildSleep |
| Child Behaviours Checklist (CBCL)^13^ | Internalising subscale | Parent | 3.95 years | MoBa |
| Child Behaviours Checklist (CBCL)^13^ | Internalising subscale | Parent | 3¾ years | POSEIDON |
| Strengths and Difficulties Questionnaire (SDQ)^18^ | Internalising subscale | Parent | 3 years | MCS |
| Strengths and Difficulties Questionnaire (SDQ)^18^ | Internalising subscale | Parent | 4 years | ALSPAC |
| Strengths and Difficulties Questionnaire (SDQ)^18^ | Emotional problems subscale | Parent | 4 years | FinnBrain |
| **Adolescence** | | | |  |
| Short Moods and Feelings Questionnaire (SMFQ)^15^ | Depressive symptoms | Parent | 11 years | MCS |
| Short Moods and Feelings Questionnaire (SMFQ)^15^ | Depressive symptoms | Parent | 13 years | ALSPAC |
| Short Moods and Feelings Questionnaire (SMFQ)^15^ | Depressive symptoms | Self | 13 years | ALSPAC |
| Strengths and Difficulties Questionnaire (SDQ)^18^ | Internalising subscale | Teacher | 11 years | MCS |
| **Adulthood** | | | |  |
| [The Clinical Interview Schedule-Revised (CIS-R)](https://www.ncbi.nlm.nih.gov/pmc/articles/PMC3347904/)^[19](https://www.ncbi.nlm.nih.gov/pmc/articles/PMC3347904/)^ | Depressive symptoms | Self | 18 years | ALSPAC |
| General Health Questionnaire (GHQ)^20^ | Depressive symptoms | Self | 18-99 (mean 47, (15)) years | GS |
| Patient Health Questionnaire-2 (PHQ2)^17^ | Depressive symptoms | Self | 21 years | MCS |

STable 2. Descriptive statistics for full sample and analytic sample by participant genetic and outcome data by age.

| **Cohorts** | **Variables** | Full sample | Analytic sample (exposure status) | | | Analytic sample (childhood emotional problems) | | | | | Analytic sample (adolescence depressive symptoms and emotional problems) | | | | | Analytic sample (adulthood depressive symptoms) | | | | |
| --- | --- | --- | --- | --- | --- | --- | --- | --- | --- | --- | --- | --- | --- | --- | --- | --- | --- | --- | --- | --- |
|  |  | *Total number* | Child PGS | Mother PGS | Father PGS | Outcome | Trio PGSes | Child PGS | Mother PGS | Father PGS | Outcome | Trio PGSes | Child PGS | Mother PGS | Father PGS | Outcome | Trio PGSes | Child PGS | Mother PGS | Father PGS |
|  |  |  | *n (%)* | *n (%)* | *n (%)* | *n = (%)* | *n (%)* | *n (%)* | *n (%)* | *n (%)* | *n (%)* | *n (%)* | *n (%)* | *n (%)* | *n (%)* | *n (%)* | *n (%)* | *n (%)* | *n (%)* | *n (%)* |
| **ALSPAC** |  |  |  |  |  |  |  |  |  |  |  |  |  |  |  |  |  |  |  |  |
|  | **Child’s sex** | | | | | | | | | | | | | | | | | | | |
|  | Male | 7695 (51.16) |  |  |  | 4916 (51.70) | 604 (53.12) | 3141 (51.38) | 2158 (49.90) | 796 (54.22) | 2502 (49.64) | 581 (52.53) | 2502 (49.64) | 1796 (48.49) | 764 (53.35) | 1993 (43.71) | 430 (48.21) | 1480 (43.85) | 1127 (43.23) | 575 (49.40) |
|  | Female | 7347 (48.84) |  |  |  | 4592 (48.30) | 533 (46.88) | 2972 (48.62) | 2167 (50.10) | 672 (45.78) | 2538 (50.36) | 525 (47.47) | 2538 (50.36) | 1908 (51.51) | 668 (46.65) | 2567 (56.29) | 462 (51.79) | 1895 (56.15) | 1480 (56.77) | 589 (50.60) |
|  | **Maternal education** | | | | | | | | | | | | | | | | | | | |
|  | Low | 7297 (62.39) |  |  |  | 5199 (59.02) | 461 (41.95) | 3189 (55.43) | 2157 (52.75) | 630 (44.49) | 2411 (52.70) | 424 (41.01) | 2411 (52.70) | 1717 (50.25) | 577 (43.68) | 2068 (51.47) | 326 (38.86) | 1509 (49.20) | 1128 (47.00) | 445 (41.20) |
|  | High | **4398 (37.61)** |  |  |  | 3610 (40.98) | **638 (58.05)** | 2564 (44.57) | 1932 (47.25) | 786 (55.51) | 2164 (47.30) | **610 (58.99)** | 2164 (47.30) | 1700 (49.75) | 744 (56.32) | 1950 (48.53) | **513 (61.14)** | 1558 (50.80) | 1272 (53.00) | 635 (58.80) |
|  | ***Continuous variables*** | *Median (IQR)* | *Median (IQR)* | *Median (IQR)* | *Median (IQR)* | *Median (IQR)* | *Median (IQR)* | *Median (IQR)* | *Median (IQR)* | *Median (IQR)* | *Median (IQR)* | *Median (IQR)* | *Median (IQR)* | *Median (IQR)* | *Median (IQR)* | *Median (IQR)* | *Median (IQR)* | *Median (IQR)* | *Median (IQR)* | *Median (IQR)* |
|  | Maternal age at birth (yrs) | 28 (6) |  |  |  | 29 (6) | 30 (6) | 29 (6) | 29 (6) | 30 (6) | 29 (6) | 30 (6) | 29 (6) | 29 (6) | 30 (6) | 29 (6) | 30 (6) | 29 (6) | 29 (6) | 30 (6) |
|  | Offspring age at outcome (yrs) | 3.95 (0.09) |  |  |  | 3.96 (0.09) | 3.95 (0.07) | 3.95 (0.08) | 3.95 (0.08 | 3.95 (0.07) | 13.75 (0.17) | 13.75 (0.17) | 13.75 (0.17) | 13.75 (0.17) | 13.75 (0.17) | 17.75 (0.33) | 17.67 (0.5) | 17.75 (0.3) | 17.67 (0.42) | 17.67 (0.5) |
| **CHILD-SLEEP** | **Sample size** | N = 1669 | - | - | - | n = 884 (52.97%) | n = 220 (13.18) | n = 773 (46.32) | n =229 (13.72) | - | - | - | - | - | - | - | - | - | - | - |
|  | **Child’s sex** | | | | | | | | | | | | | | | | | | | |
|  | Male | 882 (52.85) |  |  |  | 106 (48.18) | 402 (52.00) | 111 (48.47) | 111 (48.47) |  |  |  |  |  |  |  |  |  |  |  |
|  | Female | 787 (47.15) |  |  |  | 114 (51.82) | 371 (48.00) | 118 (51.53) | 118 (51.53) |  |  |  |  |  |  |  |  |  |  |  |
|  | **Maternal education** | | | | | | | | | | | | | | | | | | | |
|  | Low | 468 (28.73) |  |  |  | 191 (22.13) | 37 (17.21) | 167 (22.12) | 37 (16.59) |  |  |  |  |  |  |  |  |  |  |  |
|  | High | **1161 (71.27)** |  |  |  | 672 (77.87) | **178 (82.79)** | 588 (77.88) | 186 (83.41) |  |  |  |  |  |  |  |  |  |  |  |
|  | ***Continuous variables*** | *Median (IQR)* | *Median (IQR)* | *Median (IQR)* | *Median (IQR)* | *Median (IQR)* | *Median (IQR)* | *Median (IQR)* | *Median (IQR)* | *Median (IQR)* | *Median (IQR)* | *Median (IQR)* | *Median (IQR)* | *Median (IQR)* | *Median (IQR)* | *Median (IQR)* | *Median (IQR)* | *Median (IQR)* | *Median (IQR)* | *Median (IQR)* |
|  | Maternal age at birth (yrs) | 30 (6) |  |  |  | 31 (6) | 31 (6) | 30 (6) | 31 (6) |  |  |  |  |  |  |  |  |  |  |  |
|  | Offspring age at outcome (yrs) | 2.03 (0.07) |  |  |  | 2.03 (0.07) | 2.03 (0.06) | 2.03 (0.07) | 2.03 (0.06) |  |  |  |  |  |  |  |  |  |  |  |
| **Finn Brain** | **Sample size** | N = 1444 | - | - | - | N = 1444 (100) | n = 201 (13.90) | n = 1046 (72.44) | n = 223 (15.44) | n = 218 (13.06) | - | - | - | - | - | - | - | - | - | - |
|  | **Child’s sex** | | | | | | | | | | | | | | | | | | | |
|  | Male | 773 (53.53) |  |  |  | 773 (53.53) | 96 (47.76) | 557 (53.25) | 109 (48.88) | 107 (49.08) |  |  |  |  |  |  |  |  |  |  |
|  | Female | 671 (46.47) |  |  |  | 671 (46.47) | 105 (52.24) | 489 (46.75) | 114 (51.12) | 111 (50.92) |  |  |  |  |  |  |  |  |  |  |
|  | **Maternal education** | | | | | | | | | | | | | | | | | | | |
|  | Low | 392 (28.43) |  |  |  | 392 (28.43) | 54 (27.84) | 287 (28.47) | 59 (27.31) | 59 (27.96) |  |  |  |  |  |  |  |  |  |  |
|  | High | **987 (71.57)** |  |  |  | 987 (71.57) | **140 (72.16)** | 721 (71.53) | 157 (72.69) | 152 (72.04) |  |  |  |  |  |  |  |  |  |  |
|  | ***Continuous variables*** | *Median (IQR)* | *Median (IQR)* | *Median (IQR)* | *Median (IQR)* | *Median (IQR)* | *Median (IQR)* | *Median (IQR)* | *Median (IQR)* | *Median (IQR)* | *Median (IQR)* | *Median (IQR)* | *Median (IQR)* | *Median (IQR)* | *Median (IQR)* | *Median (IQR)* | *Median (IQR)* | *Median (IQR)* | *Median (IQR)* | *Median (IQR)* |
|  | Maternal age at birth (yrs) | 31 (6) |  |  |  | 31 (6) | 31 (5) | 31 (6) | 31 (6) | 31 (6) |  |  |  |  |  |  |  |  |  |  |
|  | Offspring age at outcome (yrs) | 2.04 (0.04) |  |  |  | 2.04 (0.04) | 2.04 (0.03) | 2.04 (0.04) | 2.04 (0.03) | 2.04 (0.03) |  |  |  |  |  |  |  |  |  |  |
| **MCS** | **Sample size** | N = | n = (%) | n = (%) | n = (%) | n = (%) | n = (%) | n = (%) | n = (%) | n = (%) | n = (%) | n = (%) | n = (%) | n = (%) | n = (%) | n = (%) | n = (%) | n = (%) | n = (%) | n = (%) |
|  | **Child’s sex** | | | | | | | | | | | | | | | | | | | |
|  | Male |  |  |  |  |  |  |  |  |  |  |  |  |  |  |  |  |  |  |  |
|  | Female |  |  |  |  |  |  |  |  |  |  |  |  |  |  |  |  |  |  |  |
|  | **Maternal education** | | | | | | | | | | | | | | | | | | | |
|  | Low |  |  |  |  |  |  |  |  |  |  |  |  |  |  |  |  |  |  |  |
|  | High |  |  |  |  |  |  |  |  |  |  |  |  |  |  |  |  |  |  |  |
|  | ***Continuous variables*** | *Median (IQR)* | *Median (IQR)* | *Median (IQR)* | *Median (IQR)* | *Median (IQR)* | *Median (IQR)* | *Median (IQR)* | *Median (IQR)* | *Median (IQR)* | *Median (IQR)* | *Median (IQR)* | *Median (IQR)* | *Median (IQR)* | *Median (IQR)* | *Median (IQR)* | *Median (IQR)* | *Median (IQR)* | *Median (IQR)* | *Median (IQR)* |
|  | Maternal age at birth (yrs) |  |  |  |  |  |  |  |  |  |  |  |  |  |  |  |  |  |  |  |
|  | Offspring age at outcome (yrs) |  |  |  |  |  |  |  |  |  |  |  |  |  |  |  |  |  |  |  |
| **MoBa** | **Sample size** | N = 113,967 unique (but not unrelated) children | n = 27,911 (24% of 113,967 unique children) | n = 29,961 (32% of 94,590 unique mums) | n = 28,587 (38% of 74,586 unique dads) | n = 58,072 children with observed outcome data (51% of 113,967 unique children) | n = 10,575 total trios with no relatedness between trios | n = 13,707 (unrelated) (12% of 113,967 unique (but not unrelated) children  ) | n =14,726 (unrelated) (16% of 94,590 unique (but not unrelated) mums) | n = 14,172 (unrelated) (19% of 74,586 unique (but not unrelated) dads) | - | - | - | - | - | - | - | - | - | - |
|  | **Child’s sex** | | | | | | | | | | | | | | | | | | | |
|  | Male | 58,068 (51.3%) | 14,527 (52.0%) | 15,395 (51.4%) | 14,595 (51.1%) | 29,703 (51.1%) | 5,337 (50.7%) | 7,028 (51.5%) | 7,548 (51.5%) | 7,211  (51.1%) |  |  |  |  |  |  |  |  |  |  |
|  | Female | 55,209 (48.7%) | 13,384 (48.0%) | 14,530 (48.6%) | 13,963 (48.9%) | 28,369 (48.9%) | 5,191 (49.3%) | 6,616  (48.5%) | 7,106 (48.5%) | 6,896 (48.9%) |  |  |  |  |  |  |  |  |  |  |
|  | **Maternal education** | | | | | | | | | | | | | | | | | | | |
|  | Low | 20,582 (21.0%) | 5,062 (19.5%) | 5,426 (19.7%) | 5,042 (19.1%) | 9,427 (17.2%) | 1,502 (15.0%) | 2,052 (15.9%) | 2,235 (16.1%) | 2,037 (15.2%) |  |  |  |  |  |  |  |  |  |  |
|  | High | **77,601 (79.0%)** | 20,839 (80.5%) | 22,118 (80.3%) | 21,328 (80.9%) | 45,514 (82.8%) | **8,489 (85.0%)** | 10,887 (84.1%) | 11,675 (83.9%) | 11,358 (84.8%) |  |  |  |  |  |  |  |  |  |  |
|  | ***Continuous variables*** | *Median (IQR)* | *Median (IQR)* | *Median (IQR)* | *Median (IQR)* | *Median (IQR)* | *Median (IQR)* | *Median (IQR)* | *Median (IQR)* | *Median (IQR)* | *Median (IQR)* | *Median (IQR)* | *Median (IQR)* | *Median (IQR)* | *Median (IQR)* | *Median (IQR)* | *Median (IQR)* | *Median (IQR)* | *Median (IQR)* | *Median (IQR)* |
|  | Maternal age at birth (yrs) | 30 (6) | 30 (6) | 30 (6) | 30 (6) | 30 (6) | 30 (6) | 30 (6) | 30 (6) | 30 (6) |  |  |  |  |  |  |  |  |  |  |
|  | Offspring age at outcome (yrs) | 3.08 (0.08) | 3.08 (0.08) | 3.08 (0.08) | 3.08 (0.08) | 3.08 (0.08) | 3.08 (0.08) | 3.08 (0.08) | 3.08 (0.08) | 3.08 (0.08) |  |  |  |  |  |  |  |  |  |  |
| **Poseidon** | **Sample size** | N = | n = (%) | n = (%) | n = (%) | n = (%) | n = (%) | n = (%) | n = (%) | n = (%) | n = (%) | n = (%) | n = (%) | n = (%) | n = (%) | n = (%) | n = (%) | n = (%) | n = (%) | n = (%) |
|  | **Child’s sex** | | | | | | | | | | | | | | | | | | | |
|  | Male |  |  |  |  |  |  |  |  |  |  |  |  |  |  |  |  |  |  |  |
|  | Female |  |  |  |  |  |  |  |  |  |  |  |  |  |  |  |  |  |  |  |
|  | **Maternal education** | | | | | | | | | | | | | | | | | | | |
|  | Low |  |  |  |  |  |  |  |  |  |  |  |  |  |  |  |  |  |  |  |
|  | High |  |  |  |  |  |  |  |  |  |  |  |  |  |  |  |  |  |  |  |
|  | ***Continuous variables*** | *Median (IQR)* | *Median (IQR)* | *Median (IQR)* | *Median (IQR)* | *Median (IQR)* | *Median (IQR)* | *Median (IQR)* | *Median (IQR)* | *Median (IQR)* | *Median (IQR)* | *Median (IQR)* | *Median (IQR)* | *Median (IQR)* | *Median (IQR)* | *Median (IQR)* | *Median (IQR)* | *Median (IQR)* | *Median (IQR)* | *Median (IQR)* |
|  | Maternal age at birth (yrs) |  |  |  |  |  |  |  |  |  |  |  |  |  |  |  |  |  |  |  |
|  | Offspring age at outcome (yrs) |  |  |  |  |  |  |  |  |  |  |  |  |  |  |  |  |  |  |  |
| **GS** | **Sample size** | N = 9544 (participants with a parent in the dataset) | n = 7813 (81.9% with genotype data) | n = 4094 (71.5% of 5727 mothers) | n = 2675 (46.1% of 5805 fathers) | - | - | - | - | - | - | - | - | - | - | n = 6551 (91.5%) | n = 2231 (23.4%) | n = 2231(23.4%) | n = 1387 (33.9%) | n = 1387 (51.8%) |
|  | **Child’s sex** | | | | | | | | | | | | | | | | | | | |
|  | Male | 3959 (41.5%) | 3214 (41.1%) | 2880 (41.4%) | 2012 (44.3%) |  |  |  |  |  |  |  |  |  |  | 2744 (41.9%) | 980 (43.9%) | 980 (43.9%) | 980 (43.9%) | 980 (43.9%) |
|  | Female | 5585 (58.5%) | 4599 (58.9%) | 4073 (58.6%) | 2525 (55.7%) |  |  |  |  |  |  |  |  |  |  | 3807 (58.1%) | 1251 (56.1%) | 1251 (56.1%) | 1251 (56.1%) | 1251 (56.1%) |
|  | **Maternal education** | | | | | | | | | | | | | | | | | | | |
|  | Low | 6203 (65.0%) | 5142 (65.8%) | 3987 (57.3%) | 2968 (65.4%) |  |  |  |  |  |  |  |  |  |  | 4255 (65.0%) | 1148 (48.5%) | 1148 (48.5%) | 1148 (48.5%) | 1148 (48.5%) |
|  | High | **3341 (35.0%)** | 2671 (34.2%) | 2966 (42.7%) | 1569 (34.6%) |  |  |  |  |  |  |  |  |  |  | 2296 (35.0%) | **1083 (51.5%)** | 1083 (51.5%) | 1083 (51.5%) | 1083 (51.5%) |
|  | ***Continuous variables*** | *Median (IQR)* | *Median (IQR)* | *Median (IQR)* | *Median (IQR)* | *Median (IQR)* | *Median (IQR)* | *Median (IQR)* | *Median (IQR)* | *Median (IQR)* | *Median (IQR)* | *Median (IQR)* | *Median (IQR)* | *Median (IQR)* | *Median (IQR)* | *Median (IQR)* | *Median (IQR)* | *Median (IQR)* | *Median (IQR)* | *Median (IQR)* |
|  | Maternal age at birth (yrs) | 26 (7) | 26 (7) | 26 (7) | 27 (6) |  |  |  |  |  |  |  |  |  |  | 26 (6) | 27 (6) | 27 (6) | 27 (6) | 27 (6) |
|  | Offspring age at outcome (yrs) | 33 (16) | 33 (16) | 32 (15) | 31 (13) |  |  |  |  |  |  |  |  |  |  | 32 (16) | 31 (15) | 31 (15) | 31 (15) | 31 (15) |

Notes: Maternal education was dichotomised as low, corresponding to education levels below A levels or equivalent grade in other educational systems, and high when the educational level was equal or above A level or equivalent grade.

STable 3. STROBE Statement—Checklist of items that should be included in reports of cohort studies.

|  | Item No | Recommendation | Page No |
| --- | --- | --- | --- |
| **Title and abstract** | 1 | (*a*) Indicate the study’s design with a commonly used term in the title or the abstract | 1 |
|  |  | (*b*) Provide in the abstract an informative and balanced summary of what was done and what was found | 3 |
| Introduction | | | |
| Background/rationale | 2 | Explain the scientific background and rationale for the investigation being reported | 4-5 |
| Objectives | 3 | State specific objectives, including any prespecified hypotheses | 5 |
| Methods | | | |
| Study design | 4 | Present key elements of study design early in the paper | 6 |
| Setting | 5 | Describe the setting, locations, and relevant dates, including periods of recruitment, exposure, follow-up, and data collection | 6 |
| Participants | 6 | (*a*) Give the eligibility criteria, and the sources and methods of selection of participants. Describe methods of follow-up | 7  Pg 1-7 of appendix |
|  |  | (*b*) For matched studies, give matching criteria and number of exposed and unexposed |  |
| Variables | 7 | Clearly define all outcomes, exposures, predictors, potential confounders, and effect modifiers. Give diagnostic criteria, if applicable | 7 main manuscript; 8-10 appendix |
| Data sources/ measurement | 8* | For each variable of interest, give sources of data and details of methods of assessment (measurement). Describe comparability of assessment methods if there is more than one group | na |
| Bias | 9 | Describe any efforts to address potential sources of bias | 8-9 |
| Study size | 10 | Explain how the study size was arrived at | 6 |
| Quantitative variables | 11 | Explain how quantitative variables were handled in the analyses. If applicable, describe which groupings were chosen and why | 8-9 |
| Statistical methods | 12 | (*a*) Describe all statistical methods, including those used to control for confounding | 8-9 |
|  |  | (*b*) Describe any methods used to examine subgroups and interactions |  |
|  |  | (*c*) Explain how missing data were addressed |  |
|  |  | (*d*) If applicable, explain how loss to follow-up was addressed |  |
|  |  | (*e*) Describe any sensitivity analyses |  |
| Results | | |  |
| Participants | 13* | (a) Report numbers of individuals at each stage of study—eg numbers potentially eligible, examined for eligibility, confirmed eligible, included in the study, completing follow-up, and analysed | 7, Stable 2 |
|  |  | (b) Give reasons for non-participation at each stage |  |
|  |  | (c) Consider use of a flow diagram |  |
| Descriptive data | 14* | (a) Give characteristics of study participants (eg demographic, clinical, social) and information on exposures and potential confounders | STable 2 |
|  |  | (b) Indicate number of participants with missing data for each variable of interest |  |
|  |  | (c) Summarise follow-up time (eg, average and total amount) |  |
| Outcome data | 15* | Report numbers of outcome events or summary measures over time | STable 1 |

| Main results | 16 | (*a*) Give unadjusted estimates and, if applicable, confounder-adjusted estimates and their precision (eg, 95% confidence interval). Make clear which confounders were adjusted for and why they were included | 10-11 Figures 3-6 |
| --- | --- | --- | --- |
|  |  | (*b*) Report category boundaries when continuous variables were categorized |  |
|  |  | (*c*) If relevant, consider translating estimates of relative risk into absolute risk for a meaningful time period |  |
| Other analyses | 17 | Report other analyses done—eg analyses of subgroups and interactions, and sensitivity analyses | 7-8 |
| Discussion | | | |
| Key results | 18 | Summarise key results with reference to study objectives | 10-11 |
| Limitations | 19 | Discuss limitations of the study, taking into account sources of potential bias or imprecision. Discuss both direction and magnitude of any potential bias | 14-15 |
| Interpretation | 20 | Give a cautious overall interpretation of results considering objectives, limitations, multiplicity of analyses, results from similar studies, and other relevant evidence | 12-13 |
| Generalisability | 21 | Discuss the generalisability (external validity) of the study results | 13 |
| Other information | | | |
| Funding | 22 | Give the source of funding and the role of the funders for the present study and, if applicable, for the original study on which the present article is based | 15 |

STable 4. Structural equation models disentangling the effect estimates for maternal and paternal direct, indirect, and total genetic associations on offspring emotional problems and depressive symptoms from childhood to adulthood in 7 European cohort studies.

| **Direct, Indirect, and total genetic associations by cohort study and pooled meta-analysis.** | | **Childhood** | **Adolescence** | **Adulthood** |
| --- | --- | --- | --- | --- |
|  |  | **Effect estimate (95% CI), p-value** | **Effect estimate (95% CI), p-value** | **Effect estimate (95% CI), p-value** |
| **ALSPAC** |  | **N=1,229; SDQ (4 years)** | **N=1,063; MFQ (13 years)** | **N=968; CIS-R (18years)** |
|  | **Mother** | | | |
|  | Direct effects | 0.01 (-0.02 to 0.04), p=0.567 | 0.02 (-0.02 to 0.05), p=0.281 | 0.05 (0.01 to 0.08), p=0.007 |
|  | Indirect effects | 0.02 (-0.05 to 0.09), p=0.597 | 0.09 (0.004 to 0.18), p=0.041 | -0.03 (-0.11 to 0.04), p=0.345 |
|  | Total effects | 0.03 (-0.02 to 0.08), p=0.271 | 0.11 (0.04 to 0.18), p=0.002 | 0.01 (-0.05 to 0.08), p=0.701 |
|  | **Father** | | | |
|  | Direct effects | 0.01 (-0.02 to 0.04), p=0.569 | 0.02 (-0.01 to 0.05), p=0.281 | 0.04 (0.01 to 0.08), p=0.008 |
|  | Indirect effects | -0.01 (-0.07 to 0.06), p=0.883 | -0.02 (-0.07 to 0.04), p=0.536 | -0.03 (-0.10 to 0.04), p=0.426 |
|  | Total effects | 0.004 (-0.065 to 0.073), p=0.909 | -0.001 (-0.05 to 0.05), p=0.975 | 0.01 (-0.05 to 0.08), p=0.639 |
| **Child Sleep** |  | **N=234; BITSEA (2 years)** |  |  |
|  | **Mother** | | | |
|  | Direct effects | -0.03 (-0.10 to 0.05), p=0.430 | - | - |
|  | Indirect effects | 0.08 (-0.06 to 0.23), p=0.277 | - | - |
|  | Total effects | 0.05 (-0.07 to 0.18), p=0.423 | - | - |
|  | **Father** | | | |
|  | Direct effects | -0.03 (-0.10 to 0.04), p=0.443 | - | - |
|  | Indirect effects | 0.04 (-0.11 to 0.18), p=0.621 | - | - |
|  | Total effects | 0.01 (-0.12 to 0.14), p=0.901 | - | - |
| **FinnBrain** |  | **N=172; SDQ (4 years)** |  |  |
|  | **Mother** | | | |
|  | Direct effects | 0.03 (-0.06 to 0.11), p=0.576 | - | - |
|  | Indirect effects | -0.03 (-0.19 to 0.13), p=0.720 | - | - |
|  | Total effects | -0.00 (-0.13 to 0.13), p=0.944 | - | - |
|  | **Father** | | | |
|  | Direct effects | 0.03 (-0.07 to 0.14), p=0.570 | - | - |
|  | Indirect effects | -0.05 (-0.23 to 0.13), p=0.619 | - | - |
|  | Total effects | -0.01 (-0.17 to 0.13), p=0.842 | - | - |
| **MCS** |  | **N=3,042; SDQ (3 years)** | **N=3,444; MFQ (11 years)** | **N=829; PHQ2 (18 years)** |
|  | **Mother** | | | |
|  | Direct effects | -0.02 (-0.06 to 0.02), p=307 | 0.05 (0.02 to 0.08), p=0.001 | 0.04 (-0.04 to 0.11), p=0.373 |
|  | Indirect effects | 0.09 (0.03 to 0.15), p=0.005 | -0.04 (-0.09 to 0.01), p=0.153 | 0.002 (-0.12 to 0.12), p=0.981 |
|  | Total effects | 0.07 (0.02 to 0.11), p=0.002 | 0.01 (-0.04 to 0.05), p=0.712 | 0.04 (-0.05 to 0.13), p=0.422 |
|  | **Father** | | | |
|  | Direct effects | -0.02 (-0.05 to 0.02), p=0.308 | 0.04 (0.02 to 0.07), p=0.001 | 0.04 (-0.04 to 0.11), p=0.376 |
|  | Indirect effects | 0.08 (0.02 to 0.13), p=0.011 | 0.04 (-0.01 to 0.08), p=0.117 | -0.09 (-0.19 to 0.01), p=0.090 |
|  | Total effects | 0.06 (0.01 to 0.10), p=0.017 | 0.08 (0.04 to 0.12), p=0.001 | -0.06 (-0.14 to 0.02), p=0.155 |
| **MoBa** |  | **N=10,575; CBCL (3 years)** |  |  |
|  | **Mother** | | | |
|  | Direct effects | 0.03 (0.01 to 0.06), p=0.015 | - | - |
|  | Indirect effects | 0.02 (-0.03 to 0.06), p=0.018 | - | - |
|  | Total effects | 0.05 (0.01 to 0.08), p=0.008 | - | - |
|  | **Father** | | | |
|  | Direct effects | 0.03 (0.01 to 0.06), p=0.015 | - | - |
|  | Indirect effects | -0.02 (-0.06 to 0.03), p=0.461 | - | - |
|  | Total effects | 0.01 (-0.02 to 0.05), p=0.433 | - | - |
| **Poseidon** |  | **N=223; SDQ (3 years)** |  |  |
|  | **Mother** | | | |
|  | Direct effects | -0.02 (-0.13 to 0.09), p=0.735 | - | - |
|  | Indirect effects | 0.05 (-0.19 to 0.22), p=0.561 | - | - |
|  | Total effects | 0.03 (-0.10 to 0.16), p=0.635 | - | - |
|  | **Father** | | | |
|  | Direct effects | -0.02 (-0.11 to 0.08), p=0.735 | - | - |
|  | Indirect effects | 0.13 (-0.02 to 0.28), p=0.092 | - | - |
|  | Total effects | 0.12 (-0.01 to 0.24), p=0.064 | - | - |
| **GS** |  |  |  | **N=2,413; GHQ dep (47 years)** |
|  | **Mother** | | | |
|  | Direct effects | - | - | 0.07 (0.02 to 0.12), p=0.006 |
|  | Indirect effects | - | - | 0.04 (-0.04 to 0.12), p=0.296 |
|  | Total effects | - | - | 0.11 (0.04 to 0.18), p=0.002 |
|  | **Father** | | | |
|  | Direct effects | - | - | 0.07 (0.02 to 0.12), p=0.006 |
|  | Indirect effects | - | - | -0.02 (-0.09, 0.06), p=0.694 |
|  | Total effects | - | - | 0.05 (-0.02 to 0.13), p=0.126 |
| **Pooled results** |  | **N=15,475** | **N=4,507** | **N=4,210** |
|  | **Mother** | | | |
|  | Direct effects | 0.01 (-0.01 to 0.03) | 0.04 (0.01 to 0.06) | 0.05 (0.02 to 0.08) |
|  | Indirect effects | 0.04 (0.01 to 0.07) | 0.02 (-0.11 to 0.15) | 0.00 (-0.05 to 0.05) |
|  | Total effects | 0.05 (0.02 to 0.07) | 0.06 (-0.04 to 0.15) | 0.05 (-0.01 to 0.11) |
|  | **Father** | | | |
|  | Direct effects | 0.01 (-0.01 to 0.03) | 0.03 (0.01 to 0.06) | 0.05 (0.02 to 0.07) |
|  | Indirect effects | 0.02 (-0.02 to 0.07) | 0.01 (-0.04 to 0.07) | -0.04 (-0.08 to 0.01) |
|  | Total effects | 0.03 (0.00 to 0.05) | 0.04 (-0.04 to 0.12) | 0.01 (-0.05 to 0.07) |

**References**

1.	Boyd, A. *et al.* Cohort profile: the ‘children of the 90s’—the index offspring of the Avon Longitudinal Study of Parents and Children. *Int J Epidemiol* **42**, 111–127 (2013).

2.	Fraser, A. *et al.* Cohort profile: the Avon Longitudinal Study of Parents and Children: ALSPAC mothers cohort. *Int J Epidemiol* **42**, 97–110 (2013).

3.	Northstone, K. *et al.* The Avon Longitudinal Study of Parents and Children (ALSPAC): an update on the enrolled sample of index children in 2019. *Wellcome Open Res* **4**, 51 (2019).

4.	Morales-Muñoz, I. *et al.* Impact of anxiety and depression across childhood and adolescence on adverse outcomes in young adulthood: a UK birth cohort study. *The British Journal of Psychiatry* **222**, 212–220 (2023).

5.	Smith, B. H. *et al.* Cohort Profile: Generation Scotland: Scottish Family Health Study (GS:SFHS). The study, its participants and their potential for genetic research on health and illness. *Int J Epidemiol* **42**, 689–700 (2013).

6.	Nagy, R. *et al.* Exploration of haplotype research consortium imputation for genome-wide association studies in 20,032 Generation Scotland participants. *Genome Med* **9**, 23 (2017).

7.	Fitzsimons, E. *et al.* Collection of genetic data at scale for a nationally representative population: the UK Millennium Cohort Study. *Longit Life Course Stud* **13**, 169–187 (2022).

8.	Joshi, H. & Fitzsimons, E. The UK Millennium Cohort Study: the making of a multi-purpose resource for social science and policy in the UK. *Longit Life Course Stud* **7**, 409–430 (2016).

9.	Magnus, P. *et al.* Cohort profile update: the Norwegian mother and child cohort study (MoBa). *Int J Epidemiol* **45**, 382–388 (2016).

10.	Corfield, E. C. *et al.* The Norwegian Mother, Father, and Child cohort study (MoBa) genotyping data resource: MoBaPsychGen pipeline v. 1. *BioRxiv* 2006–2022 (2022).

11.	Helgeland, Ø. *et al.* Genome-wide association study reveals dynamic role of genetic variation in infant and early childhood growth. *Nat Commun* **10**, 4448 (2019).

12.	Achenbach, T. M. & Rescorla, L. A. ASEBA preschool forms & profiles. *Burlington, VT: Research Center for Children, Youth and Families, University of Vermont* (2000).

13.	Achenbach, T. M. & Rescorla, L. A. The Achenbach system of empirically based assessment (ASEBA) for ages 1.5 to 18 years. in *The use of psychological testing for treatment planning and outcomes assessment* 179–214 (Routledge, 2014).

14.	Briggs-Gowan, M. J., Carter, A. S., Irwin, J. R., Wachtel, K. & Cicchetti, D. V. Brief Infant-Toddler Social and Emotional Assessment (BITSEA) mannual, version 2.0. (2002).

15.	Turner, N., Joinson, C., Peters, T. J., Wiles, N. & Lewis, G. Validity of the Short Mood and Feelings Questionnaire in late adolescence. *Psychol Assess* **26**, 752 (2014).

16.	Goldberg, D. P. & Hillier, V. F. A scaled version of the General Health Questionnaire. *Psychol Med* **9**, 139–145 (1979).

17.	Kroenke, K., Spitzer, R. L. & Williams, J. B. W. The Patient Health Questionnaire-2. *Med Care* **41**, 1284–1292 (2003).

18.	Goodman, R., Ford, T., Simmons, H., Gatward, R. & Meltzer, H. Using the Strengths and Difficulties Questionnaire (SDQ) to screen for child psychiatric disorders in a community sample. *The British journal of psychiatry* **177**, 534–539 (2000).

19.	Lewis, G. & Pelosi, A. J. The manual of CIS-R. *London: Institute of Psychiatry* (1992).

20.	Goldberg, P. The detection of psychiatric illness by questionnaire. *Maudsley monograph* (1972).
